# Exploration of Alzheimer’s Disease MRI Biomarkers Using *APOE4* Carrier Status in the UK Biobank

**DOI:** 10.1101/2021.09.09.21263324

**Authors:** Jingnan Du, Zhaowen Liu, Lindsay C. Hanford, Kevin M. Anderson, Jianfeng Feng, Tian Ge, Randy L. Buckner

## Abstract

Large-scale datasets enable novel strategies to refine and discover relations among biomarkers of disease. Here 30,863 individuals ages 44-82 from the UK Biobank were analyzed to explore MRI biomarkers associated with Alzheimer’s disease (AD) genetic risk as contrast to general effects of aging. Individuals homozygotic for the E4 variant of *apolipoprotein E* (*APOE4*) overlapped non-carriers in their 50s but demonstrated neurodegenerative effects on the hippocampal system beginning in the seventh decade (reduced hippocampal volume, entorhinal thickness, and hippocampal cingulum integrity). Phenome-wide exploration further nominated the posterior thalamic radiation (PTR) as having a strong effect, as well as multiple diffusion MRI (dMRI) and white matter measures consistent with vascular dysfunction. Effects on the hippocampal system and white matter could be dissociated in the homozygotic *APOE4* carriers supporting separation between AD and cerebral amyloid angiopathy (CAA) patterns. These results suggest new ways to combine and interrogate measures of neurodegeneration.

## Introduction

Alzheimer’s Disease (AD) is a progressive neurodegenerative illness that is the most common cause of dementia in older adults. While post-mortem pathology remains the gold-standard for diagnosis, advances in *in vivo* research staging have demonstrated the importance of biomarkers that can detect disease-specific pathology (ß-amyloid and tau) as well as effects of neurodegeneration (e.g., hippocampal atrophy) (Jack et al., 2010; Johnson et al., 2012; Jack et al., 2013). In addition to their value for classification (e.g., Jack et al., 2016), measures of neurodegeneration track disease progression (Dubois et al., 2014) and thus may play a critical role in measuring response to treatment and clarifying disease-modifying factors.

Various measures of the hippocampal formation are the most consistently relied on biomarkers of neurodegeneration associated with AD including age-adjusted cross-sectional and longitudinal MRI measures of volume (e.g., Jack et al., 1998; Mielke et al., 2012; Schuff et al., 2009). Diffusion within the hippocampal cingulum (CgH) shows a parallel effect (Salat et al., 2010; Wang et al., 2012; Nir et al., 2013; Rieckmann et al., 2016). Broader cortical atrophy is also detected by structural MRI using volumetric registration of longitudinal data and thickness estimates in cross-sectional contrasts (Scahill et al., 2002; Buckner et al., 2005; Dickerson et al., 2009; Fennema-Notestine et al., 2009; Gordon et al., 2018). These broader cortical regions also demonstrate low metabolism measured by [18F]-flurodeoxyglucose-PET (FDG-PET) (Herholz et al., 2002; Buckner et al., 2005; Jagust et al., 2010).

Here we took a novel approach to identify and prioritize neurodegenerative biomarkers for AD. We utilized the vast power of the large UK Biobank (UKBB) neuroimaging dataset to screen for MRI variables that differed based on genetic risk of AD, complementing recent approaches that have explored aging more broadly (e.g., Smith et al., 2020). The UKBB is a large prospective population-based cohort that recruited approximately 500,000 middle-aged to older community volunteers with a sub-sample of participants contributing brain MRI (Littlejohns et al., 2020). In the UKBB sample, 30,000 participants are expected to develop AD dementia by 2027 (Sudlow et al., 2015). Genetic information is also available including information about whether individuals carry one or two copies of the E4 variant of *apolipoprotein E* (*APOE4*). *APOE4* is a common allele that is consistently associated with increased risk of AD (Corder et al., 1993; Strittmatter et al., 1993). Homozygotic carriers of *APOE4*, who are infrequent in the general population, are substantially more likely to express memory decline and dementia than non-carriers (Roses, 1996; Caselli et al., 2009; Qian et al., 2017). *APOE4* is also a strong genetic risk for vascular amyloid, described as cerebral amyloid angiopathy (CAA), which may be associated with overlapping or distinct MRI biomarker patterns. Given its scale, the UKBB dataset has a large number of individuals who are homozygotic carriers of the *APOE4* allele.

Our analyses began with an initial pool of 42,184 individuals who had MRI data and then culled the sample down to 30,863 high-quality datasets. Within that sample we asked the question of which MRI measures differed as a function of *APOE4* carrier status. As will be shown, expected measures that are commonly used for measuring AD-related neurodegeneration emerged at the top of the screen, including hippocampal volume. In addition, novel measures emerged that have to date been underrepresented in the field’s exploration of AD neurodegenerative biomarkers, some of which may reflect pathophysiological mechanisms associated with CAA.

## Results

### Control measures display well-behaved age and sex effects

A total of 30,863 individuals remained for analyses after the participant selection (Fig. 1; female: 53.0%, age: 64.0±7.5y, age range = 44.6 – 81.8y). Initial analyses focused on control measures to establish the validity of our approach. The cross-sectional estimated total intracranial volume (eTIV), global cortical thickness and Freesurfer-derived bilateral hippocampal volume are displayed for seven age bins (44-49y, 50-54y, 55-59y, 60-64y, 65-69y, 70-74y, 75-82y) split by genetically defined sex in Fig. 2. We specifically chose to plot the data by age bins so that the details of the variance and scatter can be visualized for each age cohort, as well as to allow the stability of the data to be easily appreciated, since each age bin represents data that are independent from the other age bins.

**Fig. 1.**
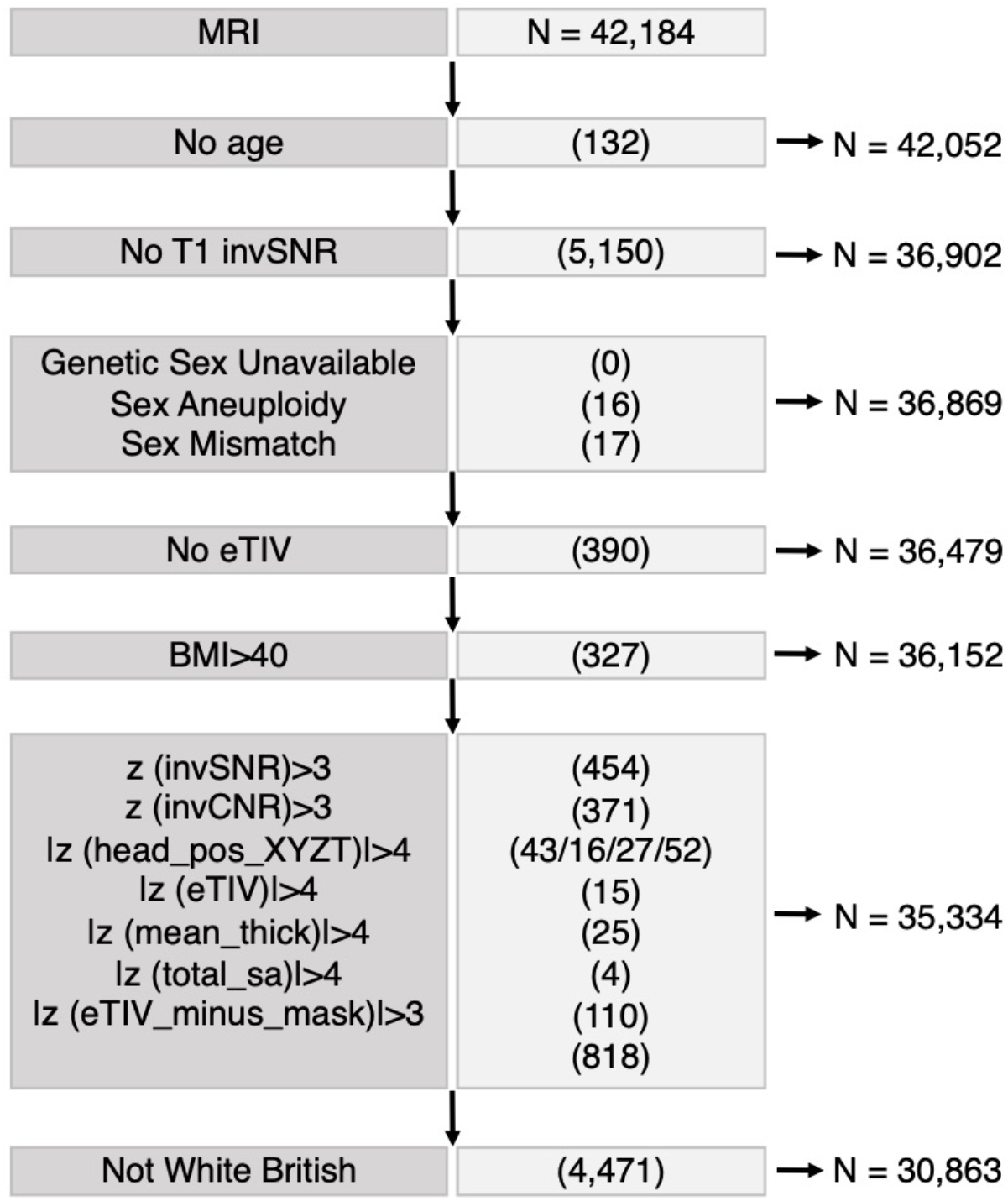
Flowchart of the participant selection. A flow chart illustrates the selection of participants for the present analyses. Starting from N = 42,184 participants, selection criteria yielded a final sample of N=30,863 used for MRI analyses stratified by *APOE4* carrier status. eTIV = estimated total intracranial volume, invSNR = inverted signal-to-noise ratio in T1, invCNR = inverted contrast-to-noise ratio in T1, mean_thick = mean global cortical thickness, total_sa = total surface area, eTIV_minus_mask = volume-ratio of MaskVol-to-eTIV in the whole brain generated by subcortical volumetric segmentation (ASEG), BMI = body mass index, head_position_XYZT includes four independent variables: scanner lateral (X) brain position, scanner transverse (Y) brain position, scanner longitudinal (Z) brain position and scanner table (T) position.

**Fig. 2.**
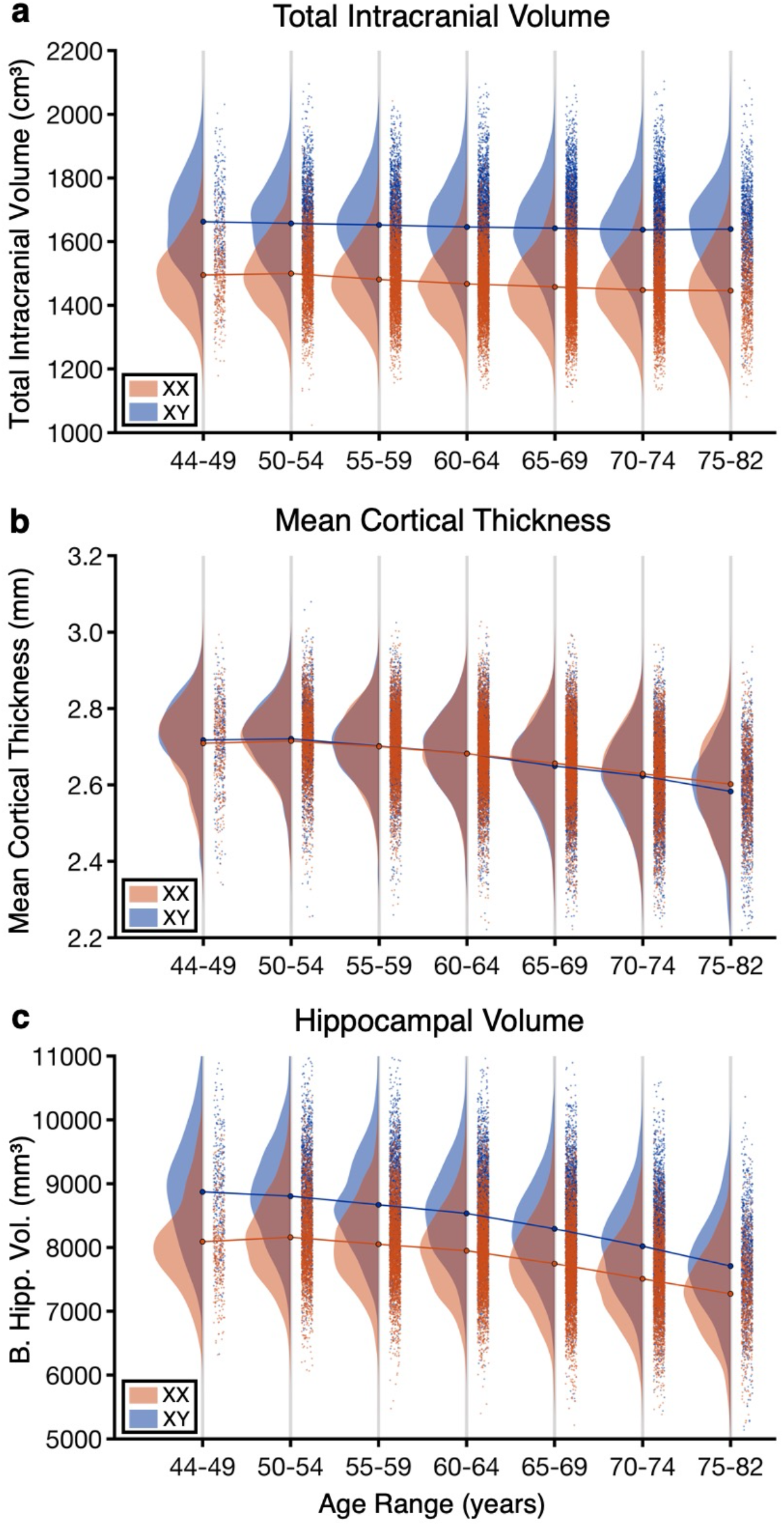
Analyses of control brain measures suggest data stability and validity. Each data point represents the value of the brain measure for a single individual. The data distributions are shown separately for men (XX) in blue and women (XY) in red for each age bin. A blue curve connects the mean values of the distributions of all the age bins for men and a red curve for women. **(a)** Estimated total intracranial volume (eTIV) by age and sex. **(b)** Mean cortical thickness by age and sex. **(c)** Bilateral hippocampal volume by age and sex.

eTIV, which is a proxy for head size, displayed a minimal effect of age for both men and women consistent with the expectation that head size differs minimally with age (Davis and Wright, 1977; Buckner et al., 2004). The effect of sex, by contrast, was robust for every age bin. Overall, men displayed a 12.0% larger eTIV than women with the effect nearly the same for each of the seven age bins (range = 10.4% to 13.3%).

Mean cortical thickness is known to reduce with age with minimal or no sex difference in young adults. Fig. 2b confirms this effect in the present sample. Over the age span examined, there is a 4.5% reduction in cortical thickness. In the earlier age bins men and women have almost identical estimates. In the oldest individuals, there is slightly reduced thickness in men as compared to women, consistent with a sex difference in the progression of age effects.

Bilateral hippocampal volume demonstrates both effects of age and sex (Fig. 2c). The sex difference in volume is robust and present in all age bins, paralleling the head-size difference. A robust volume decrease is noted with age with a steeper decrease in men than in women (age x sex interaction, t(30,862)= −9.17, p <0.001).

These collective results support that the UKBB brain measures are stable and robust to known effects of age and sex, with minimal bias of registration across the age span.

### APOE4 carrier status predicts differences in known AD neurodegenerative biomarkers

To explore the effect of *APOE4* on imaging measures, we split the sample by *APOE4* carrier status in each age bin. Among the 30,863 subjects, there were 22,158 (71.8%) carrying no APOE4 allele, 7,996 (25.9%) carrying one copy, and 709 (2.3%) carrying two copies. Thus the large sample size of the UKBB allowed for a sizeable sample of homozygotic *APOE4*/*APOE4* (*E4/E4*) carriers despite the rarity in the population. Given that there were only 19 *E4/E4* carriers in the 44-49y age bin and 40 in the 75-82y age bin, we focused all further analyses on the middle five age bins which respectively included 87, 119, 148, 189 and 107 *E4/E4* carriers (see Table 1).

**Table 1.** Sample size of groups stratified by *APOE4* allele carrier status and age.

| Age Range<br>*APOE4* Status | 44-49 | 50-54 | 55-59 | 60-64 | 65-69 | 70-74 | 75-82 |
| --- | --- | --- | --- | --- | --- | --- | --- |
| 0 (C/C) | 477 | 2644 | 3524 | 4478 | 5271 | 4282 | 1482 |
| 1 (C/T) | 191 | 1011 | 1347 | 1679 | 1895 | 1385 | 488 |
| 2 (T/T) | 19 | 87 | 119 | 148 | 189 | 107 | 40 |
| Percentage of<br>2 (T/T) | 2.77% | 2.32% | 2.38% | 2.35% | 2.57% | 1.85% | 1.99% |

Covariate-adjusted bilateral hippocampal volume was significantly lower in the oldest groups of E4/E4 carriers as contrast to the individuals with no copies (Fig. 3a; p<0.05 in 60-64y, p<0.01 in 65-69y, p<0.001 in 70-74y). Similarly, we explored whether diffusion tensor imaging (DTI) and neurite orientation dispersion and density imaging (NODDI) estimates of bilateral CgH were affected. The intracellular volume fraction (ICVF) dMRI parameter was selected a priori, but as will be shown later, the observed effects generalized across many dMRI parameters. Fig. 3b reveals the robust carrier status effect on the CgH in older individuals (p<0.01 in 60-64y, p<0.001 in 65-69y, p<0.001 in 70-74y).

**Fig. 3.**
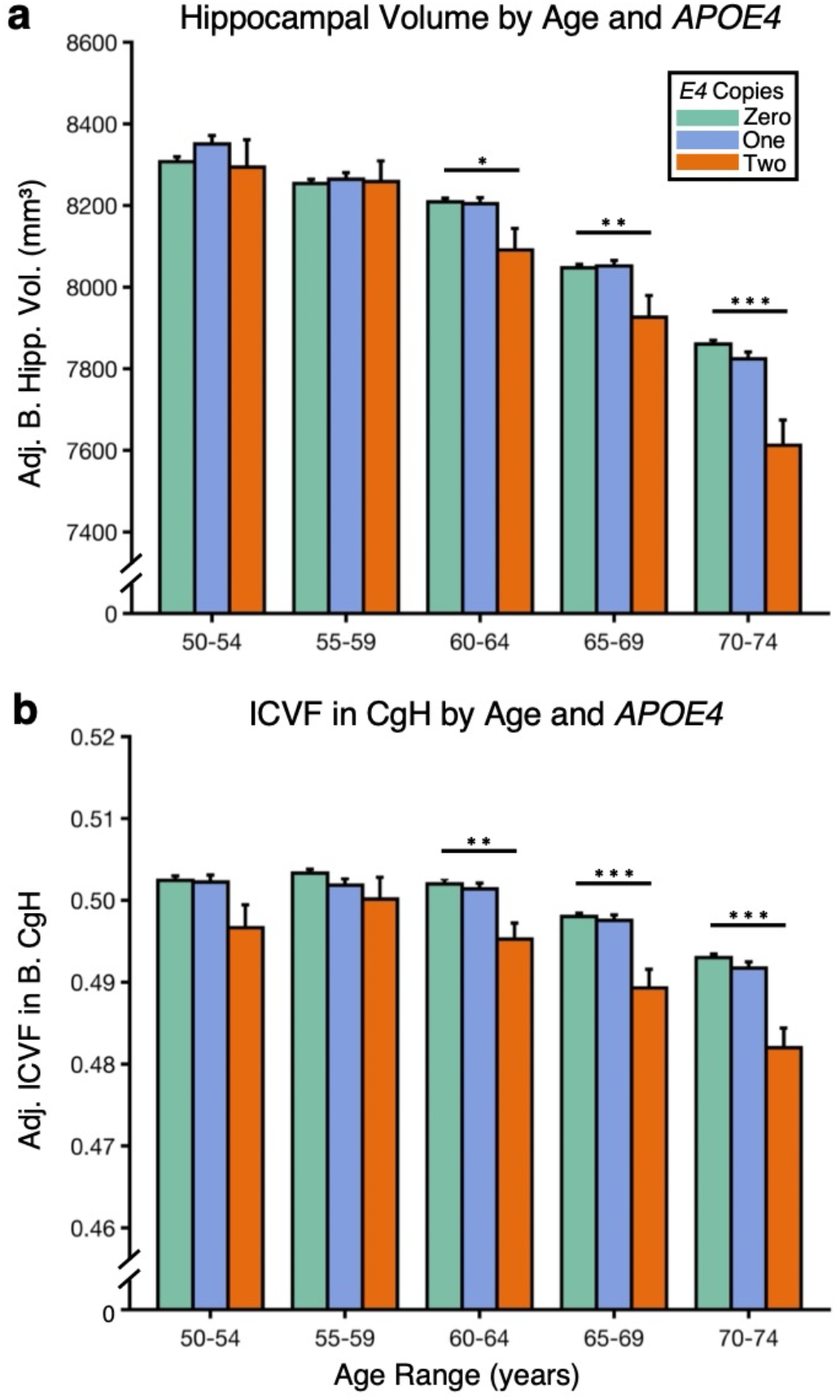
*APOE4* carrier status predicts differences in established MRI measures of neurodegeneration. Measures are plotted for the five age bins with sufficient samples of homozygotic *APOE4* allele carriers. Each age bin is separated into groups of individuals carrying zero copies of *APOE4*, one copy, or two copies. **(a)** Covariate-adjusted bilateral hippocampal volume is plotted. Note that there is no effect of carrier status in the early age bins, while a growing effect emerges in the older bins with *E4/E4* carriers showing reduced hippocampal volume, consistent with early-stage AD neurodegeneration. The effect is independently significant in each of the three oldest age bins. **(b)** Covariate-adjusted dMRI measurement of the hippocampal cingulum (using ICVF) are similarly plotted. Again the effect is independently significant in each of the three oldest age bins. Note that the dMRI data are acquired from a scan separate from the structural volume data. Error bars indicate standard error of the mean. * represents p<0.05; ** represents p<0.01; *** represents p<0.001. CgH = hippocampal cingulum.

These results reveal that stratifying the sample by genetic risk manifests an age-dependent pattern of biomarker differences that parallels known effects of AD neurodegeneration. Thus, genetic risk, in this sample, might serve as a screening variable for biomarkers of AD neurodegeneration.

To further explore the signal properties of stratifying by E4/E4 carrier status, we conducted additional analyses on these well-established AD neurodegenerative biomarkers. For each measure, we first regressed age and age2, and the same covariates as in our other analyses (see Methods) to explore the stability of the measures within our model. The reason age and age2 were regressed here is to detect individuals who have extreme values above and beyond differences expected by age. We then replotted the data by age bin to illustrate the probability of an individual having a measure that falls within the lowest 5% of the data. In a sample where all individuals are drawn from the same population, the estimates will be flat hovering around 0.05 for all age bins and groups. In a sample where some individuals had a heightened probability of an extreme value, such as we predict for *E4/E4* carriers, their probabilities will be selectively higher. Finally, to extend the analyses beyond a single threshold we examined a continuous Q-Q plot where the obtained probability of each measure was plotted across different thresholds from 5% to 95% for the oldest age bin.

Fig. 4 shows the results for hippocampal volume. With age, age^2^, and covariates regressed, hippocampal volume is well behaved and flat with a normal distribution of values (Fig. 4a). Individuals from the *E4/E4* groups in the older age bins (60-74y) are more likely to have a low hippocampal volume than would be expected by chance (Fig. 4b). Individuals with no or one *APOE4* copy reveal a well-behaved distribution with their probabilities of a low volume hovering around the chance-expected 0.05 level across age bins. Furthermore, the obtained probability of having a low hippocampal volume in the *E4/E4* group in the oldest age bin (70-74y) is higher than the other groups across thresholds, consistent with a shift in the overall distribution of values (Fig. 4c). Fig. 5 illustrates the results for CgH, derived from dMRI, which parallel the effects from the T_1_ hippocampal volumes, but with the effect being most clear in the two oldest age bins (65-74y).

**Fig. 4.**
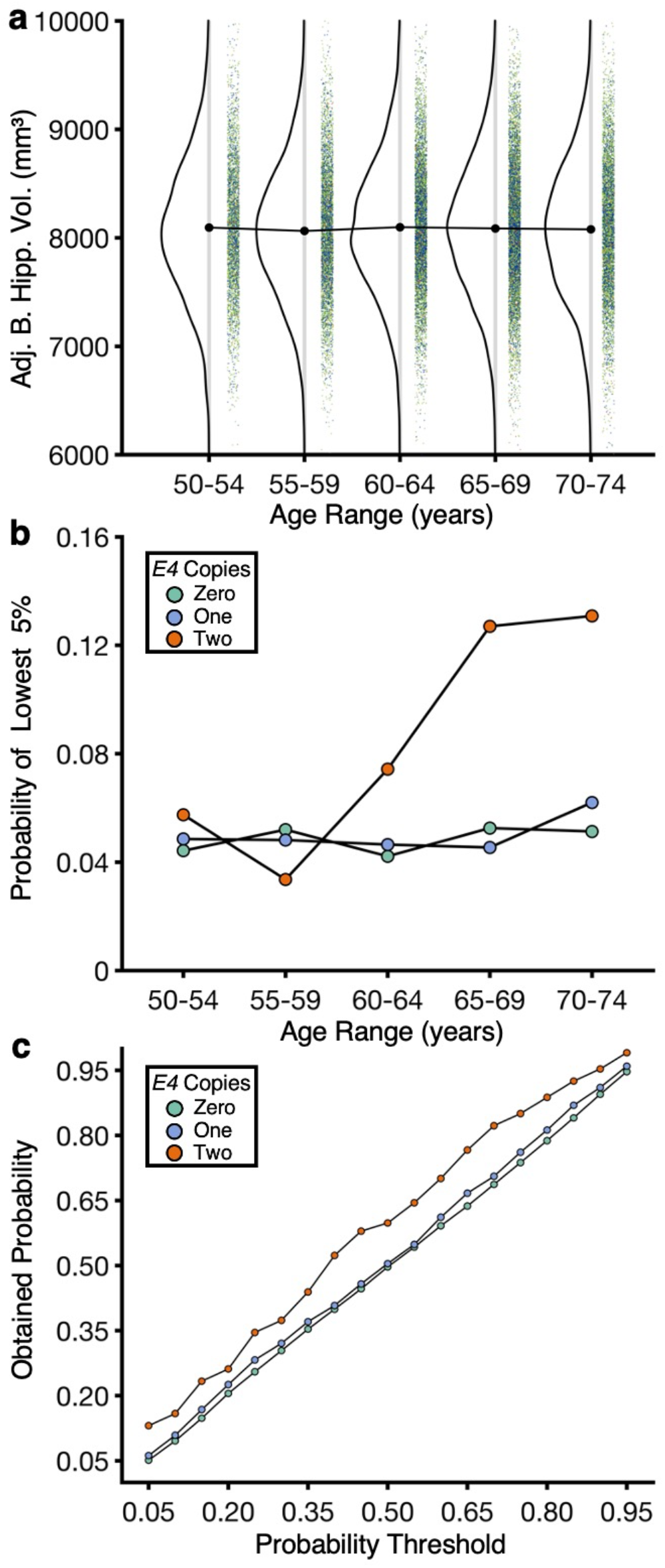
Further exploration of *APOE4* carrier status on hippocampal volume. **(a)** The distributions of the bilateral hippocampal volumes across age bins are illustrated after regression of covariates. Note that the measure is well behaved and flat with a normal distribution of values. (**b)** The obtained probability that individuals will possess a low hippocampal volume (bottom 5 percent of the whole sample cut-off) as a function of *APOE4* carrier status. **(c)** The obtained probabilities against different cut-off probability thresholds in the 70-74y age bin.

**Fig. 5.**
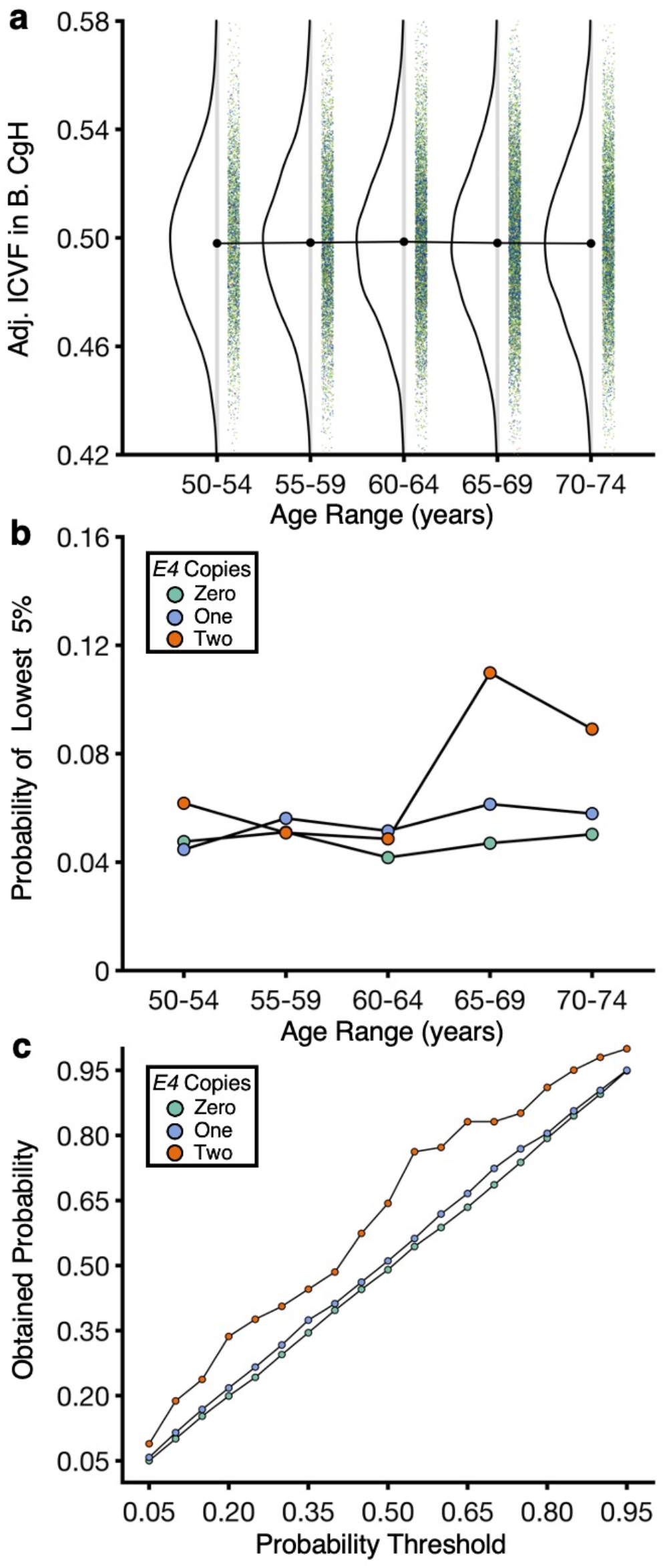
Further exploration of *APOE4* carrier status on the hippocampal cingulum. **(a)** The distributions of the bilateral hippocampal cingulum (estimated from the dMRI data) across age bins are illustrated after regression of covariates. Paralleling the effects for hippocampal volume, the measure is well behaved and flat with a normal distribution of values. **(b)** The obtained probability that individuals with possess low ICVF in the hippocampal cingulum (bottom 5 percent of the whole sample cut-off) as a function of *APOE4* carrier status. **(c)** The obtained probabilities against different cut-off probability thresholds in the 70-74y age bin.

Taken collectively these results for established AD neurodegenerative biomarkers reveal that stratification by *E4/E4* carrier status in the UKBB data yields robust effects in the older age bins. Given this result, the next set of explorations generalized this observation by quantifying and plotting the effect of *E4/E4* carrier status for many of the available MRI biomarkers in the UKBB as a screen to identify and prioritize candidate AD neurodegenerative biomarkers.

### *APOE4* carrier status reveals numerous candidate AD neurodegenerative biomarkers

A total of 689 IDPs were examined including 55 T_1_ subcortical volumetric measures, 202 cortical measures of regional thickness, volume and surface area, and 432 diffusion MRI skeleton measures (48 white matter tract labels x 9 types of dMRI measures for each label). For each T_1_ structural and dMRI imaging measure, the difference between the homozygotic *E4/E4* carrier group and the age-matched group with no *APOE4* copies was estimated. Specifically, the descriptive statistic Cohen’s *d* was used to measure the group differences in those individuals ages 65-74y (based on the results in Figs. 3-5). Within this age range, which provided the most signal in the earlier analyses, there were 296 *E4/E4* carriers and 3,280 individuals with no copies of *APOE4. Post hoc* analyses exploring a broader age range (60–74y) minimally changed the results.

As expected given the originating analyses and the literature, hippocampal volumes and multiple diffusion parameters associated with the CgH showed robust effects (Fig. 6). Of note, the effects generalized across related structures within the hippocampal diencephalon and its cortical projection zones, as well as across related measures of the same structure. For example, within the thickness measures, the entorhinal cortex (EC) revealed a moderate effect size. Within the diffusion measures, the CgH showed a robust effect for the ICVF measure as used in the earlier analyses but also effects in fractional anisotropy (FA) as well as mean diffusivity (MD, in the expected, opposite direction).

**Fig. 6.**
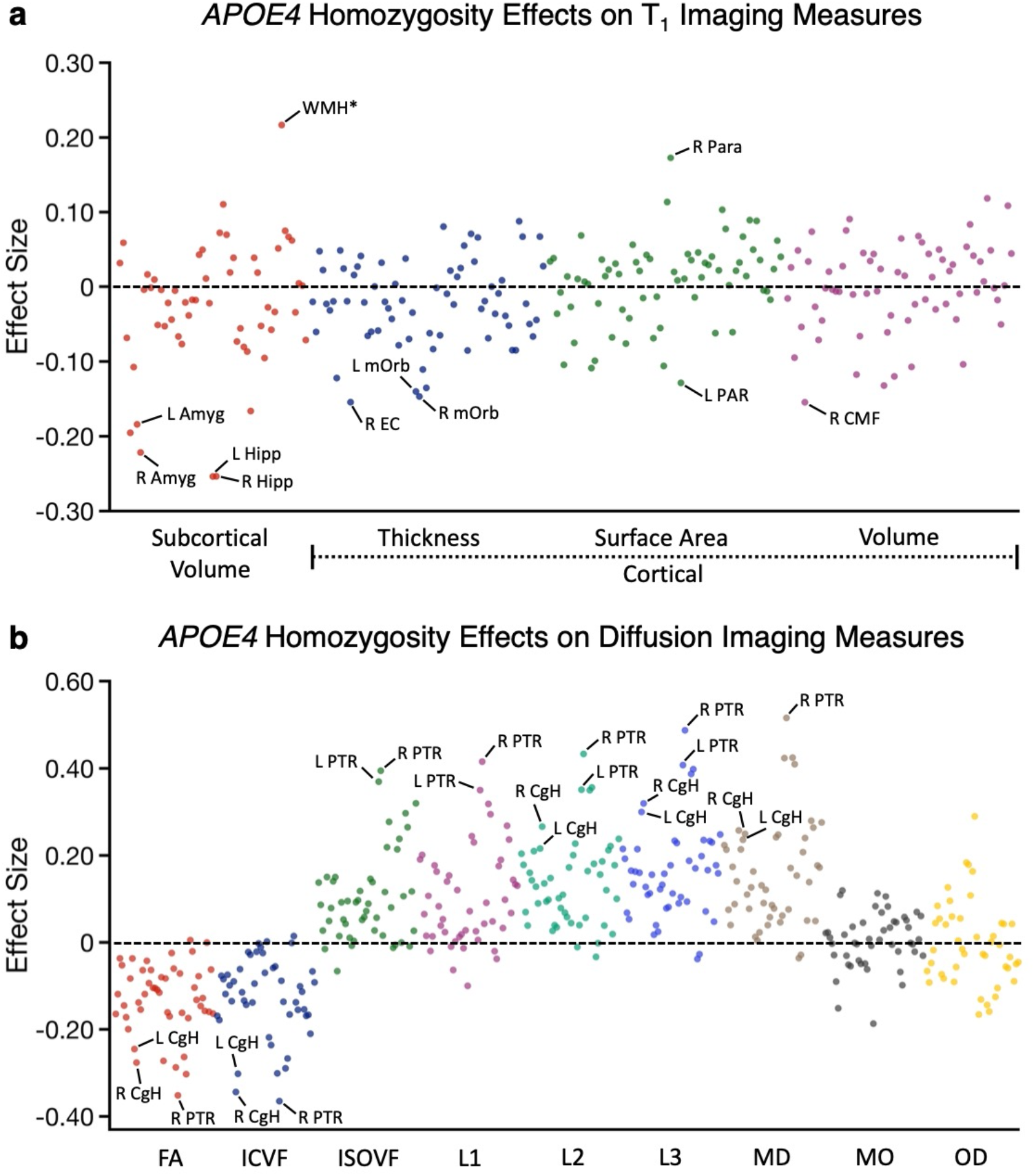
Data-driven exploration of candidate neurodegenerative biomarkers from T_1_ structural and dMRI measures. **(a)** The effect size of being a homozygotic *APOE4* carrier on each T_1_ structural measure is plotted. Estimates were obtained from individuals ages 65-74y. The Y-axis labels illustrate the class of measure with the color of the dots corresponding to the labelled class below. A subset of the measures showing large effect size is labeled, such as hippocampal volume and entorhinal cortex. **(b)** The effect size of being a homozygotic *APOE4* carrier on each dMRI measure is plotted with class of measure noted in the Y-axis separated by color. Amyg = amygdala; Hipp = hippocampal volume; WMH* = total volume of white matter hypointensities; EC = entorhinal cortex; mOrb = medial orbitofrontal cortex; PAR = pars opercularis cortex; Para = paracentral cortex; CMF = caudal middlefrontal cortex; CgH = hippocampal cingulum; PTR = posterior thalamic radiation.

Beyond these effects, there were many other measures that were not our *a priori* targets that revealed clear signal. These included bilateral amygdala volumes, estimates of white matter hypointensities (WMH*), and a robust dMRI effect in the posterior thalamic radiation (PTR) across multiple diffusion parameters. We refer to white matter hypointensities as WMH* because the measure is solely based on quantification in the T_1_ image, not the more traditional approach using a FLAIR (Fluid-attenuated inversion recovery) image. The UKBB dataset also has a measure that reflects the traditional FLAIR quantification strategy (as used in Alfaro-Almagro et al., 2018). The two measures (T_1_-based WMH* and the traditional quantification with FLAIR) are correlated at *r* = 0.85, and the *APOE4* effect sizes are nearly the same for this analysis: 0.217 for WMH* as used here and 0.216 for the measure that uses the FLAIR image. The complete list of measures screened and their estimated effect sizes can be found in Supplementary Tables S1 and S2. Of particular interest, the effect within the PTR was larger than the dMRI effects in the CgH or the T_1_ volume effects in the hippocampus.

To further appreciate the spatial topography of the effects, the effect sizes for all of the cortical surface volume differences and the dMRI tract differences (using ICVF) are plotted in Fig. 7. In the instance of the cortical surface, bilateral regions were combined and plotted together on the same hemisphere. These resulting spatial patterns yield several notable findings.

**Fig. 7.**
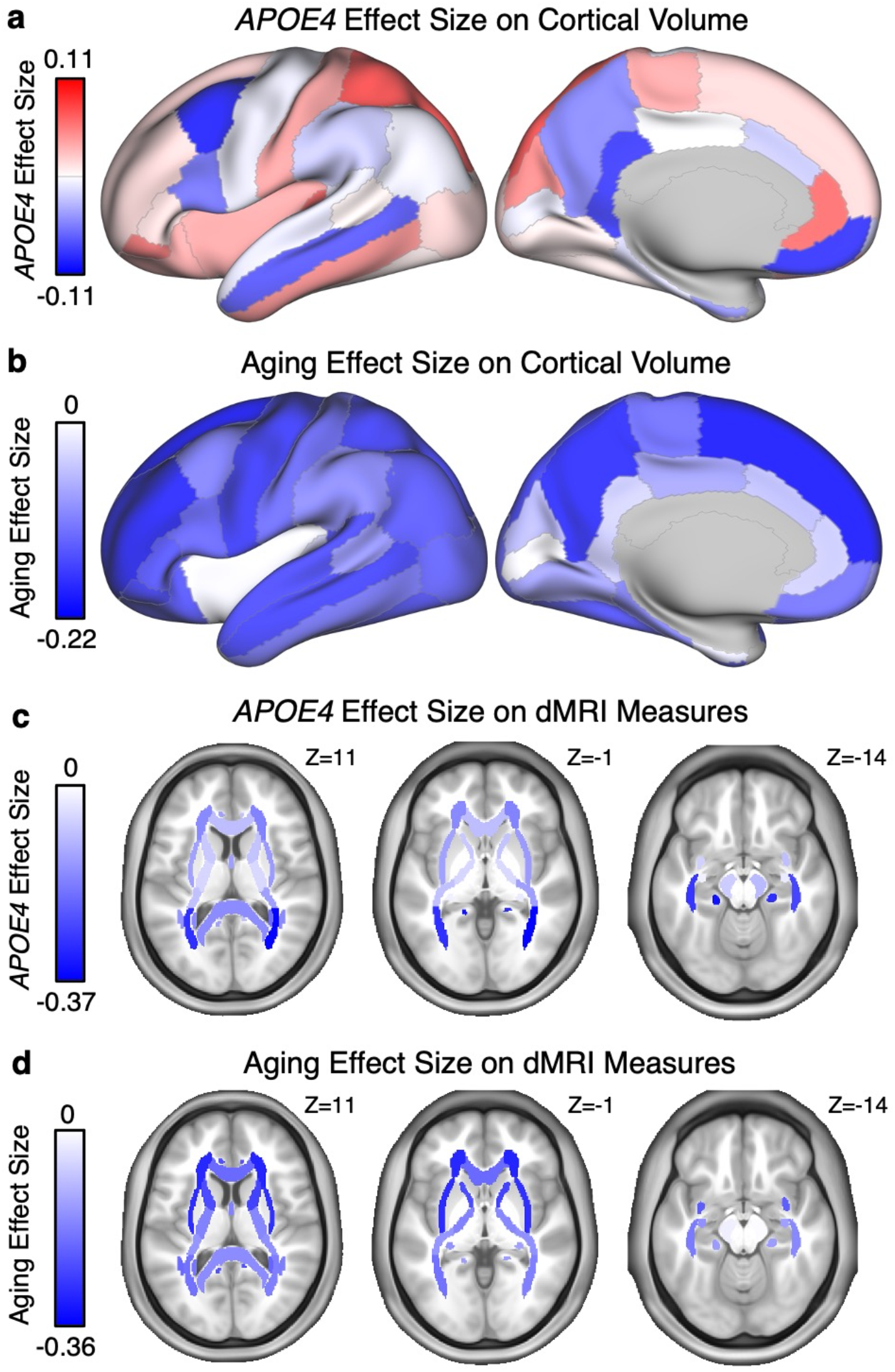
Spatial topography of effect sizes for *APOE4* carrier status and aging. **(a)** The effect size of being a homozygotic *APOE4* carrier is plotted on the cortical surface for each T_1_ cortical estimate of volume. The views represent the lateral and medial surface, and the colors represent the effects sizes with dark blue showing the greatest carrier status reduction in volume. The plotted data come from bilateral regions. Note the distributed *APOE4* carrier effect pattern includes posterior and anterior midline regions, and posterior parietal cortex, regions reliably implicated as affected in early stages of AD. **(b)** The effect size of aging is plotted on the same cortical surface map for individuals carrying zero copies of *APOE4*. **(c)** The effect size of being a homozygotic *APOE4* carrier is plotted for dMRI measures of ICVF within each separate tract. Note the tracts with the largest effect sizes are located in posterior regions. **(d)** The effect size of aging for the same dMRI measures. Note that aging shows larger effect sizes for anterior brain regions.

First, the cortical topography of the *E4/E4* effect on the cortical surface is distributed across multiple regions in a pattern consistent with prior examinations of the natural history of AD. *Relatively* strong cortical effects were evident along the posterior and anterior midline, extending into the precuneus and retrosplenial cortex, as well as along temporal association cortex and the posterior parietal cortex. Thus, while the individual cortical regions did not achieve as strong effect sizes as some of the subcortical volumes (e.g., hippocampus), the pattern of preferential effects is consistent with the known neurodegenerative pattern in early-stage AD (e.g., Scahill et al., 2002; Buckner et al., 2005; Dickerson et al., 2009; Fennema-Notestine et al., 2009; Gordon et al., 2018). Second, the dMRI effect pattern is preferentially posterior that, as will be discussed, is consistent with CAA and distinct from the anterior-preferential effect pattern common to typical aging.

### Age effects reveal a distinct biomarker landscape

The results above reveal effects of *E4/E4* carrier status within the oldest individuals, with age regressed. To contextualize these effects and contrast them with age effects, we conducted a parallel analysis in the sample of individuals carrying no copies of *APOE4* across the full age range (N=22,158). The aging effect size of each imaging phenotype was estimated as the Pearson’s product moment correlation with age. Fig. 8 illustrates the results.

**Fig. 8.**
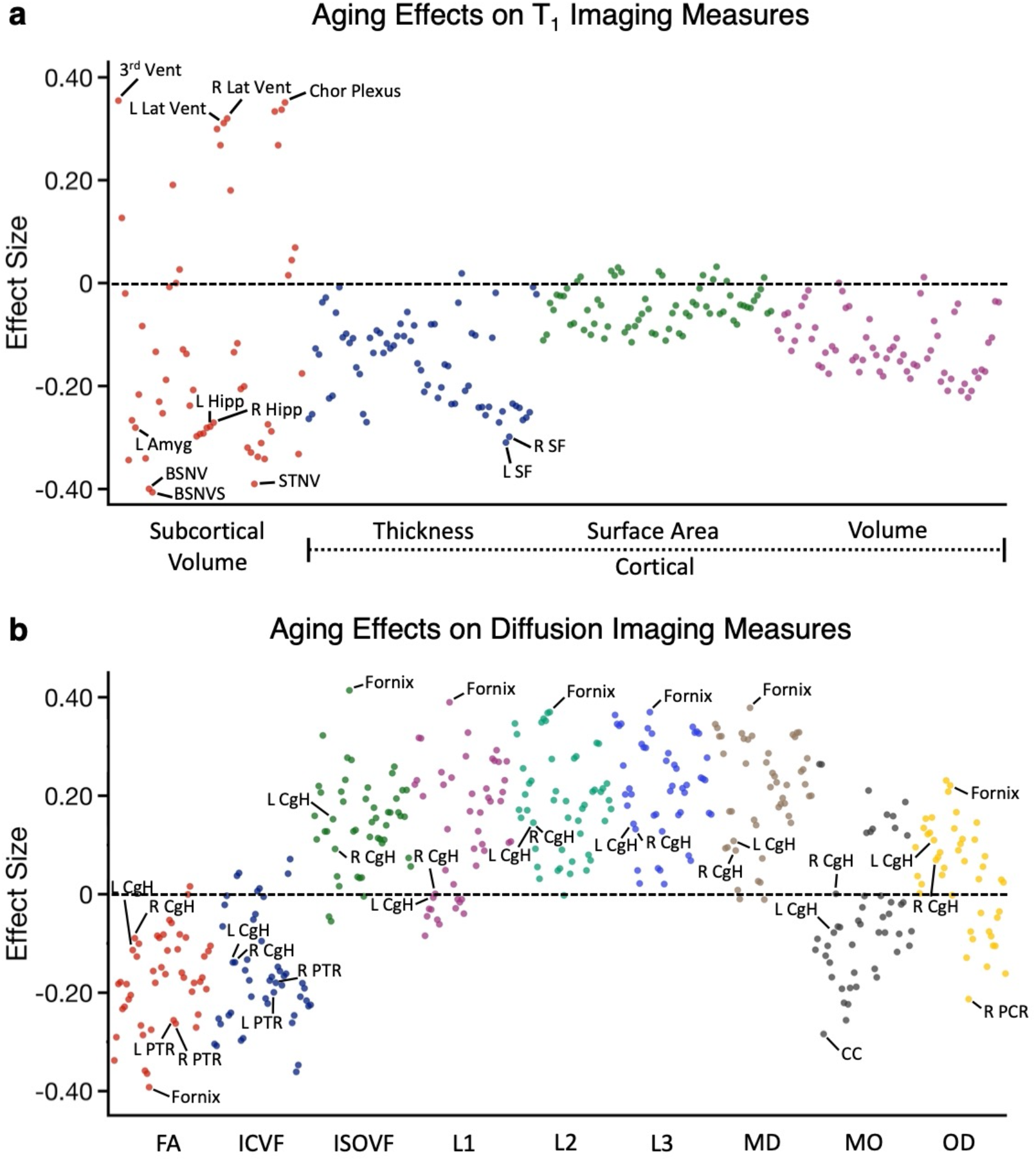
Data-driven exploration of aging effects from T_1_ structural and dMRI imaging measures. **(a)** The effect size of aging on each T_1_ structural measure is plotted. Estimates were obtained from all the subjects with no copies of *APOE4*. The Y-axis labels the class of measure with the color of the dots corresponding to the labelled class below. A subset of the measures showing large effect size were labeled, such as fornix, 3^rd^ ventricle, BrainSegNotVentSurf. **(b)** The effect size of aging on each dMRI measure is plotted with class of measure noted in the Y-axis and separated by color. Amyg: amygdala; 3^rd^ Vent: 3^rd^ ventricle; BSNVS: BrainSegNotVentSurf; BSNV: BrainSegNotVent; Lat Vent: Lateral Ventricle; STNV: SupraTentorialNotVent; Hipp: hippocampal; Chor Plexus: choroid-plexus; SF: superior frontal; CgH: hippocampal cingulum; PTR: posterior thalamic radiation; CC: body of corpus callosum; PCR: posterior corona radiata.

While some of the measures that showed effects of *E4/E4* carrier status also showed age effects (e.g., hippocampal volume), they mostly did not stand out. A broad range of T_1_ structural measures showed strong age effects. Several proxies for global whole brain differences revealed particularly strong age effects. For example, the whole brain segmentations that excluded the ventricles showed strong negative effects (BrainSegNotVentSurf, BSNVS and BrainSegNotVent, BSNV) and ventricle volumes showed strong positive effects (3^rd^ ventricle and lateral ventricles). These effects presumably reflect age-dependent reduction of gray and white matter volume and parallel increases in ventricle / cerebral spinal fluid (CSF) spaces, or direct effects of increasing CSF pressure with age. Of further note, specific measures of the hippocampal diencephalon showed strong age effects including multiple dMRI measures of the fornix.

The spatial topography of the aging effects were plotted in the same manner as the effects of A*POE4* carrier status allowing their distinct patterns to be compared (Fig. 7b and 7d). Note that these are relative differences, as the effect size estimates for *APOE4* carrier status and aging were based on analyses that differed in power.

### Effects of AD genetic risk can stand out against effects of aging

To contrast the effects of *E4/E4* carrier status and the effects of aging, the two sets of estimates were plotted against one another for both the T_1_ structural and dMRI imaging measures in Fig. 9. There was a low to modest correlation between the two (r^2^ = 0.12 for the T_1_ imaging measures and r^2^ = 0.43 for the dMRI measures). What is informative in terms of AD neurodegenerative biomarker development is that several measures showed strong carrier status effects that were large relative to their expected effect sizes based on age. These included hippocampal volume and WMH* in the T_1_ imaging measures and CgH and PTR among the dMRI measures. For example, carrying two copies of *APOE4* allele reduces the bilateral hippocampal volume by 165.92 mm^3^ in the older group ages 65-74y, which is equivalent to 6.68 years of the effect of normal aging in individuals carrying zero copies of *APOE4*.

**Fig. 9.**
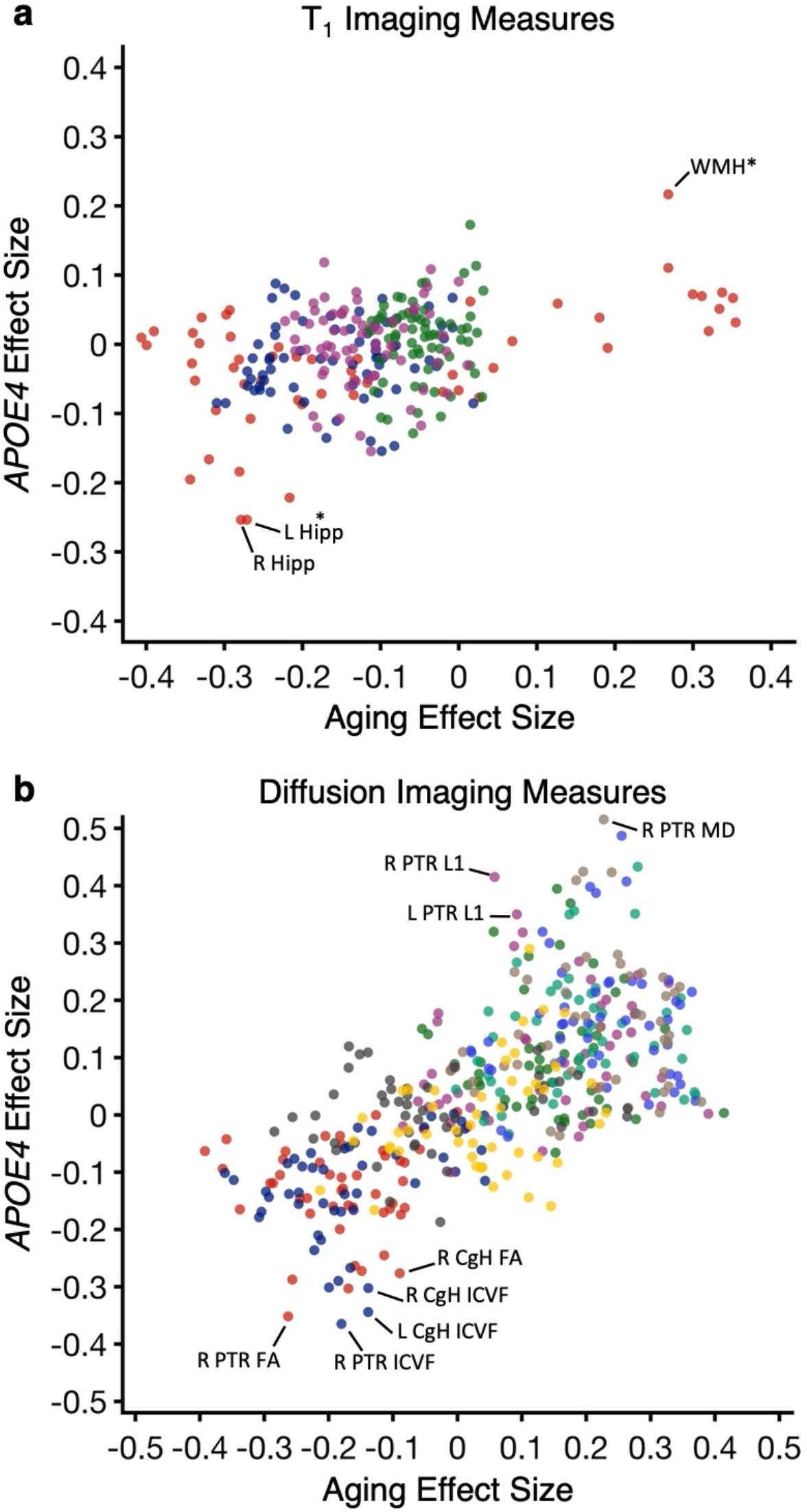
*APOE4* carrier status reveals disproportionate effects on measures beyond aging. **(a)** The effect size of being a homozygotic *APOE4* carrier was plotted directly against the effect size of aging for each T_1_ structural measure. A subset of the measures that show large AD effect sizes disproportionate to their aging effects are labeled, such as hippocampal volume and WMH*. **(b)** The effect size of being a homozygotic *APOE4* carrier against effect size of aging on each dMRI structural measure is plotted. A subset of the measures that show disproportionately large AD effect sizes are again labeled, such as ICVF and FA in hippocampal cingulum, and ICVF, FA, L1 and MD in posterior thalamic radiation. Hipp = hippocampal volume; WMH* = total volume of white matter hypointensities; CgH = hippocampal cingulum; PTR = posterior thalamic radiation; ICVF: intra-cellular volume fraction; FA: fractional anisotropy; MD: mean diffusivity.

### Dissociation between hippocampal system and broader white matter effects

An unexpected result was the strong effects on both traditional MRI measures of neurodegeneration of the hippocampal system as well as effects that may reflect vascular or blood-brain barrier compromise. The posterior pattern of the white matter effects as revealed in the dMRI data is consistent with CAA. As CAA is common in AD-type neurodegeneration, there has long been debate about the relation between the two pathological effects (e.g., Vinters, 1992). Given this background we conducted a *post hoc* set of analyses to explore whether the two classes of effect are associated or dissociable.

The specific question we asked was whether individuals with evidence of neurodegeneration in the hippocampal system were also the same individuals with evidence of white matter neurodegeneration in the PTR. We began by selecting the 274 homozygotic *APOE4* carriers (22 of the 296 *E4/E4* carriers did not have preprocessed dMRI measures) ages 65-74y. To this point all analyses considered these individuals as a group so the present explorations, which examine variation within the group, are orthogonal and unbiased. Within this high risk genetic group, we asked if variation in bilateral hippocampal volume is associated with one or both of bilateral CgH ICVF and bilateral PTR ICVF. We similarly asked if WMH* is associated with one or both of bilateral CgH ICVF and bilateral PTR ICVF. These measures reflect the strongest effects in our earlier explorations that mark these two classes of effect (see Figs. 6, 8, and 9). All covariates were regressed as in earlier analyses including age and age^2^ so as to isolate the effects of neurodegeneration. For the dMRI measures, we also regressed an average of three frontal ICVF measures (genu of corpus callosum and bilateral anterior corona radiata) to remove global between-individual differences in dMRI measurement quantities. The results are shown in Fig. 10.

**Fig. 10.**
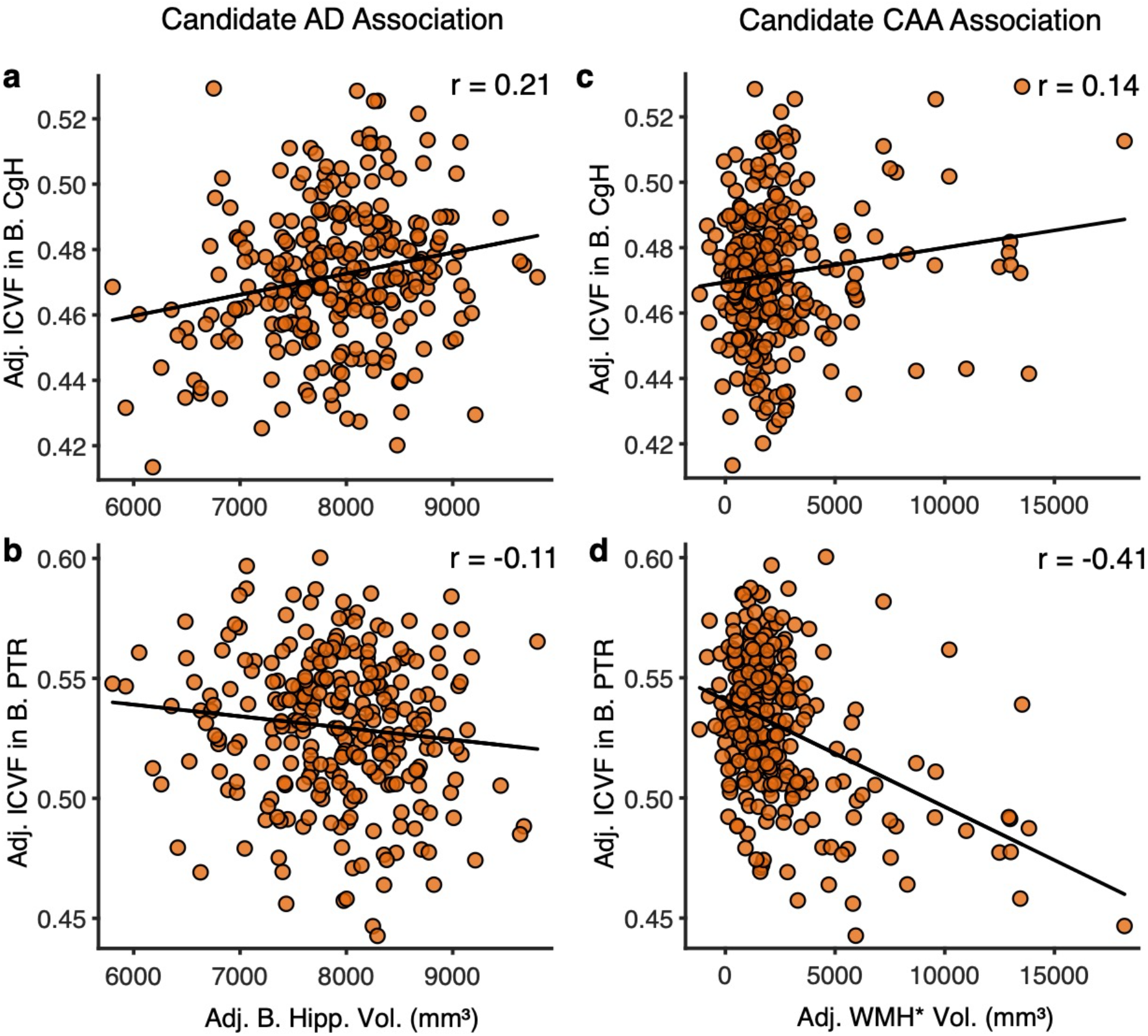
Homozygotic *APOE4* carriers demonstrate separable hippocampal system and posterior white matter effects. Simple correlations are plotted that are chosen to emphasize biomarkers differentially linked to AD (anchoring from differences in hippocampal volume) and CAA (anchoring from differences in WMH*). Candidate AD associations are plotted to the left (and predicted to be positive) while candidate CAA associations are plotted to the right (and predicted to be negative. **(a)** The correlation between adjusted bilateral hippocampal volume and adjusted ICVF in bilateral CgH. **(b)** The correlation between adjusted bilateral hippocampal volumes and adjusted ICVF in bilateral PTR. **(c)** The correlation between adjusted WMH* volume and adjusted ICVF in bilateral CgH. **(d)** The correlation between adjusted bilateral hippocampal volumes and adjusted ICVF in bilateral PTR. Note that the two dMRI measures are differentially correlated with hippocampal volume and WMH* suggesting a dissociation. WMH* = white matter hypointensities measured via the T_1_ structural image; CgH = hippocampal cingulum; PTR = posterior thalamic radiation; ICVF = intra-cellular volume fraction.

Adjusted bilateral hippocampal volumes positively predicted dMRI measurement of the CgH (Fig. 10a). Individuals who show small hippocampal volumes tend to be those that, on average, show reduced integrity of the CgH as measured by dMRI (r = 0.21, p<0.001). No such positive association was noted for dMRI measurement of the PTR (Fig. 10b; r = −0.11). By contrast, T_1_ measurement of WMH* showed a negative correlation with dMRI measurement of the PTR (Fig. 10d; r = −0.41, p<0.001). This pattern suggests that the greater the extent of WMH*, the lower the integrity of the PTR as measured by dMRI. No such effect was noted for hippocampal volume (Fig. 10c; r = 0.14). This pattern is consistent with a dissociation between the *APOE4* effects on the hippocampal system and the broad effects on white matter. While these two effects are likely present to varying degrees across individuals, the separation observed here indicates the effects do not closely track one another. Rather, they can be dissociated. Note further that this dissociation directly crosses measurement acquisition modality: the T_1_ hippocampal volume estimate predicted a dMRI estimate within the CgH, and the T_1_ measurement of WMH* predicted a dMRI estimate of the white matter posterior fiber pathway (the PTR).

## Discussion

Neuroimaging and genetic data from the large UKBB cohort were analyzed to identify and compare neurodegenerative MRI biomarkers based on *APOE4* carrier status. Three findings emerged. First, results confirmed the importance of well-utilized MRI biomarkers (T_1_ structural measurement of hippocampal volume) and added weight to previously proposed but less utilized biomarkers (dMRI measurement of the CgH). Second, numerous candidate biomarkers emerged that have strong effects but have not been prominent to date in MRI measurement of AD neurodegeneration (Figs. 6 and 8 and Supplementary Tables 1 and 2). A broad class of these, including multiple dMRI measures with emphasis on posterior regions (e.g., PTR) and global WMH*, may reflect vascular or blood-brain barrier dysfunction associated with CAA. Third, the strategy assigned an effect size for each MRI phenotype as well as an estimate of the effect of aging, enabling the possibility of combining and weighting measures to maximize sensitivity to progression of neurodegeneration. We discuss these results as well as limitations of our approach.

### Use of APOE4 carrier status to identify and prioritize candidate biomarkers

To appreciate the utility and limitations of the biomarker screening strategy taken here it is important to consider the approach in the context of other strategies that have been used to identify and prioritize neurodegenerative MRI biomarkers in AD. Numerous efforts, including those undertaken by some of the present authors, have used the natural history of AD progression as a means to stage the disease and follow the progression of biomarkers. The Alzheimer’s Disease Neuroimaging Initiative (ADNI) and affiliated international projects have been paradigmatic examples of this approach, suggesting the utility of specific MRI biomarkers (e.g., Fennema-Nostine et al., 2009; Schuff et al., 2009; Nir et al., 2013), as well as contributing data to frame the relationships and longitudinal trajectories among biomarkers (e.g., Jack et al., 2010; 2013). Convergent natural history approaches have used population-based cohorts of sporadic AD (Jack et al., 2016) as well as family-based studies of individuals with autosomal dominant mutations that convey exceptional risk for developing AD (Bateman et al., 2012). These collective studies are the foundation of the field.

In this paper, we adopt a complimentary screening strategy to identify biomarkers tied to genetic Alzheimer’s risk in the general population. A vast general-utility dataset was able to be harnessed to identify and prioritize MRI biomarkers in an unbiased and data-driven manner. For those biomarkers known to the field, a relative weighting is suggested, with hippocampal volume, as expected, emerging among the top T_1_ structural measures. For others, including measures using dMRI, the present screen suggests weightings and identification of new measures that extend beyond current priorities of the field.

Although the utility of dMRI measures to track AD neurodegeneration, especially for the CgH, has precedent (e.g., Salat et al., 2010; Wang et al., 2012; Nir et al., 2013; Rieckmann et al., 2016), this modality has not yet become a priority for the field. In some large-scale studies, and virtually all clinical trials to our knowledge, dMRI has been underemphasized or not acquired. The present quantitative results of *APOE4* effect sizes across MRI measures reprioritize dMRI measures. The effect size for the CgH was larger than that noted for hippocampal volume. This result, by itself, suggests that dMRI measures of the hippocampal formation can act as a convergent, independent measure of AD neurodegeneration. The correlation of the T_1_ estimates of hippocampal volume and dMRI measures of the CgH in the *E4/E4* carriers further supports measuring the CgH to track AD-type neurodegeneration (Fig. 10).

A barrier to considering dMRI measures as biomarkers is the impression that the acquisition will be long and disproportionately sensitive to motion, in part because a trend of the field is to acquire high angular resolution, multi-shell acquisitions to resolve crossing fibers, such as utilized by the Human Connectome Project (Sotiropoulos et al., 2013; Fan et al., 2014). The CgH is a largely monodirectional bundle (see Mori et al., 2005; see also Wu et al., 2016) that affords itself to measurement from single-shell acquisitions that have relatively low angular resolution (e.g., 15-20 directions). It may well be possible to acquire relevant, quantitatively stable dMRI in 90s or less with available acquisitions and potentially in under a minute as dMRI acquisitions adopt newer acceleration strategies (e.g., compressed sensing acceleration; Tobisch et al., 2018; Mussard et al., 2020).

### Expansion of the traditional AD biomarker emphasis

While several of the identified biomarkers are consistent with neurodegeneration of the hippocampal system, including multiple measures arising from distinct MRI acquisitions and analysis approaches (e.g., hippocampal volume, entorhinal cortex thickness, white matter properties of the CgH), many of the identified measures are much broader and capture effects that likely arise from vascular or blood-brain barrier dysfunction. The data support this in several ways. Among the T_1_ imaging measures, a particularly strong effect was found for WMH*. WMH* are non-specific effects linked to vascular compromise. Similarly, within the dMRI measures, while the CgH shows a large effect size, it is notable that many other dMRI measures also display strong effects, with the measures of FA and ICVF having numerous negative effects across anatomically distinct fiber tracts, and MD revealing the expected mirrored positive effects in the same tracts (Fig. 6b).

AD is associated with vascular amyloid deposits often described as CAA (Vinters, 1992; Serrano-Pozo et al., 2011; Liu et al., 2013; DeTure and Dickson, 2019). Post-mortem examination of the brains of *E4/E4* carriers has specifically suggested stronger-than-expected vascular amyloid deposition in relation to AD disease severity (as measured by Braak staging) (Hultman et al., 2013), and individuals with familial amyloid precursor protein (APP) duplication show substantial vascular amyloid deposits (Rovelet-Lecrux et al., 2006). Longitudinal neuroimaging assessments of individuals with heterogenous forms of dominantly inherited AD reveal increased prevalence of cerebral microhemorrhages (e.g., Joseph-Mathurin et al., 2021). Providing a candidate mechanistic explanation for these effects, recent explorations of early-stage AD suggest *APOE4* carrier status conveys risk for dysfunction of the blood-brain barrier (Montagne et al., 2020).

Of further note, CAA preferentially affects posterior parietal and occipital regions (Serrano-Pozo et al., 2011) and spatial clustering of hemorrhages measured with MRI in individuals with probable CAA reveals a preferentially posterior distribution (Rosand et al., 2005). It is thus of interest that the strongest dMRI measure identified by our screen was the PTR. Fig. 7 illustrates the anatomical distribution of the full set of dMRI effects. While the effect of aging reveals an anterior-biased pattern, the effects of *APOE4* carrier status are preferentially posterior, consistent with CAA.

Most broadly, the numerous effects in MRI biomarkers that likely reflect vascular dysfunction add to the growing literature highlighting the interplay between vascular compromise and traditional pathological emphases on parenchymal (non-vascular) amyloid deposition. Our results open avenues for development of disease-tracking MRI biomarkers and also weigh in on the importance of exploring how AD pathophysiology affects the hippocampal system specifically but also vascular or blood-brain barrier dysfunction typically associated with CAA. Both are identified in our screen for MRI biomarkers associated with *APOE4* homozygosity. Our *post hoc* analyses that examined the correlation between hippocampal system effects against broader white matter effects within the group of 274 *E4/E4* carriers revealed that the two classes of effect can be dissociated. WMH* associated with dMRI effects in the PTR (Fig. 10d) while hippocampal volume associated with dMRI effects in the CgH (Fig. 10a). This intriguing result, which cuts across MRI acquisition modalities to target distinct neurodegenerative sequalae, suggests that the individuals presenting with a CAA-type neurodegenerative pattern can be distinct from those with the classical AD-type neurodegenerative pattern. Thus, while *APOE4* is a strong risk for both cascades, the neurodegenerative patterns are not expressed to the same degree within each individual.

### Limitations

The present results are based on a cross-sectional analysis of a specific genetic risk factor for AD. While longitudinal analyses often converge on analyses that examine between-group differences, there are known pitfalls. Our approach will weight higher MRI biomarkers that do not have unaccounted for between-subject variance at baseline. Head-size is a strong predictor of hippocampal volume at baseline, and can be regressed, to accommodate a major component of between-subject anatomical variation. By contrast, cortical gyrification is more variable and less controlled in our analyses. Longitudinal designs, because each individual serves as her or his own control, are less sensitive to between-individual differences. Thus, some important markers may be underemphasized in our cross-sectional screen. As one example, within-individual longitudinal analysis in sporadic AD (Scahill et al., 2002; Buckner et al., 2004) and dominantly inherited AD (Benzinger et al., 2013; Gordon et al., 2018) have both converged on accelerated atrophy of the precuneus / posterior cingulate. While these effects can be detected in cross-sectional measurements (e.g., Dickerson et al., 2009; Fennema-Notestine et al., 2009), they do not differentiate themselves as clearly. In our analyses, the expected cortical atrophy pattern is detected consistent with the prior literature, including effects along the posterior midline extending into retrosplenial cortex (Fig. 7a). However, these effects are not particularly strong. Our design has its greatest value in its ability to positively nominate candidate biomarkers in a data-driven manner, including those that have not previously been a focus. The approach lacks the ability to measure MRI biomarkers that depend on longitudinal assessment (e.g., the boundary shift integral; Freeborough and Fox, 1997).

A future direction for this research will be to expand the genetic analyses beyond *APOE4*. Though *APOE4* genetic risk is a powerful means to identify effects of AD-type neurodegeneration, genetic risk for AD extends well beyond the locus that includes *APOE4. APOE4* is a specific risk factor associated with increased amyloid accumulation and, as shown by the data patterns, has two related but dissociable sets of effects, one linked more to traditional AD-type neurodegeneration of the hippocampal system and a second associated with CAA emphasizing posterior vascular effects. Moving forward, the present framework can be extended to explore more diverse genetic targets. For example, the approach can be generalized to other common variations of relative large effect size, including the top hits in recent genome-wide association studies (GWAS)(Harold et al., 2009, Kunkle et al., 2019; Jansen et al. 2019, Lambert et al., 2013). As another example, the polygenic risk score (PRS; Wray et al., 2021), which aggregates genetic risk from hundreds to thousands of potential contributing single-nucleotide polymorphisms, could also add explainable variance related to the biological processes of AD.

### Conclusions

The vast power of the large UKBB dataset was harnessed to explore effects of *APOE4* carrier status on MRI brain measures. Results identified well-established AD MRI biomarkers and revealed strong effects across multiple measures that target the hippocampal system, across MRI acquisition types and analyses. In addition, there were widespread effects across measures of white matter that are consistent with vascular compromise. By using an unbiased data-driven approach, all of these measures are assigned effect-size weights that can be used for future targeted studies that focus on developing sensitive measures to track neurodegeneration as well as motivate mechanistic hypotheses about the singular or multiple cascades that are present in AD.

## Methods

### Overview

Neuroimaging data from the UKBB were analyzed to identify measures that progressively differed by *APOE4* carrier status as a function of age. Analyses proceeded in four stages. (1) Initial control analyses verified that UKBB measures display known age-dependent patterns, independent of AD genetic risk. For these initial analyses, eTIV, mean cortical thickness, and Freesurfer-derived hippocampal volume were examined because they have predictable, distinct effects by age and sex. (2) The next analyses focused on established measures of AD neurodegeneration including the volume of the hippocampus and diffusion properties within the CgH, to determine if *APOE4* carrier status would reveal differences. For these analyses, the effects of *APOE4* carrier status were tracked across five five-year age bins (50-74y) that each contained sufficient numbers of participants carrying two copies of *APOE4*. As shown, *APOE4* carrier status revealed robust effects that emerged in the older age bins but not the younger age bins. (3) Enabled by the demonstration that *APOE4* carrier status yields robust effects on known measures, the screen was generalized to extensive T_1_ structural imaging and dMRI measures. For each measure, the effect of *APOE4* carrier status was examined controlling for age, and the effect of age was separately examined after removing the *APOE4* carriers, thus allowing AD effects to be directly contrast with effects of aging. (4) Motivated by the results, the data were replotted to reveal disassociations between effects of aging and *APOE4* carrier status, and between biomarker patterns linked to traditional AD-type neurodegeneration versus those potentially associated with CAA.

### UKBB Cohort

The UKBB (http://www.ukbiobank.ac.uk) is a large prospective population-based cohort study that recruited approximately 500,000 community volunteers recruited between 2006–2010, living close to multiple assessment centers in England, Scotland, and Wales (Sudlow et al., 2015; Miller et al., 2016). Participants provided comprehensive assessments including demographics, lifestyle, and disease history, with linkages to electronic medical records at baseline. Since 2014, a subset of participants underwent brain MRI scans (Littlejohns et al., 2020) with 42,184 available at the time of our analysis. At the time of their scans, the included participants were ages 44-82y. UKBB received ethical approval from the North West Centre Research Ethics Committee (REC number 11/NW/0382). The current analyses were conducted under the approved UKBB application 32568.

### MRI acquisition and processing

Details of the MRI protocol and processing are publicly available (Miller et al., 2016; Alfaro-Almagro et al., 2018). Briefly, all the brain imaging data were acquired using 3T Siemens Skyra System (Siemens, Erlangen, Germany) with the VD13A SP4 operating system and the vendor-supplied 32-channel head coil. The UKBB data includes six MRI modalities. We used two of the six including T_1_-weighted structural and diffusion MRI. T1-weighted imaging was performed by a three-dimensional magnetization-prepared acquisition with gradient echo sequence (TI = 880ms, TR = 2,000ms, TE = 2.01ms, flip angle = 8°, resolution = 1.0×1.0×1.0 mm). T_1_ structural images were used to derive measures related to volumes, cortical thickness and morphology of brain structures. dMRI data were acquired using an echo-planer, single-shot Stejskal-Tanner pulse sequence. 36 sections (resolution = 2.0×2.0×2.0 mm) in 50 distinct diffusion-weighted directions were acquired with two b-values (b = 1000 and 2000 sec/mm^2^). Tensor fitting utilizes b = 1000 s/mm^2^ data to calculate FA, axial diffusivity (AD), radial diffusivity (RD), and MD maps. In addition to DTI fitting, the diffusion images were fed into NODDI modelling (Zhang et al., 2012) to generate parameters including ICVF, ISOVF (isotropic or free water volume fraction) and OD (orientation dispersion index). Tract-Based Spatial Statistics (TBSS) measures were then calculated by averaging the skeletonized images of each dMRI map within a set of 48 standard-space tract masks defined by the Johns Hopkins University White Matter Atlas (JHU ICBM-DTI-81space) (Mori et al., 2005 and Mori et al., 2007).

### Brain imaging data and image-derived phenotypes (IDPs)

Brain imaging data were processed through an automated image processing pipeline (Alfaro-Almagro et al., 2018) by the UKBB imaging team to create image-derived phenotypes (IDPs) (https://www.fmrib.ox.ac.uk/ukbiobank/fbp). In this study, 689 separate IDPs were included in analysis that spanned global measures (e.g., eTIV, whole brain volume, mean cortical thickness, global surface area, total WMH*), regional volumes (e.g., hippocampus, amygdala), regional measures of cortical thickness, volume and surface area, and dMRI measures of local tracts. dMRI measures include DTI and NODDI parameters: FA; ICVF; ISOVF; L1; L2; L3; MD; MO; OD.

### UKBB genetics

UKBB genetic data quality control was performed based on the metrics provided by the UKBB. Specifically, we removed individuals flagged for genotype missingness and heterozygosity outliers, with putative sex chromosome aneuploidy, had mismatches between genetically inferred sex and self-reported sex, and were excluded from kinship inference or autosome phasing. We retained unrelated individuals who were self-identified and genetically confirmed white British. To estimate *APOE4* carrier status, dosage data for the single nucleotide polymorphism (SNP) rs429358 (chr19:44908684; T/C) was extracted from genome-wide data in BGEN format.

### Data screening

Prior to analyses, a participant selection procedure was applied (Fig. 1). In addition to the genetic exclusions above, individuals with no age recorded were removed. Individuals with no T_1_ inverted signal-to-noise ratio were removed. Individuals with no eTIV estimation output in the Freesurfer pipeline were excluded. Individuals with BMI higher than 40 were removed. Additionally, extreme values of the distribution of inverted signal-to-noise (SNR) ratio in T_1_, inverted contrast-to-noise ratio (CNR) in T_1_, mean global cortical thickness, total surface area, volume-ratio of MaskVol-to-eTIV in the whole brain generated by subcortical volumetric segmentation, scanner lateral (X) brain position, scanner transverse (Y) brain position, scanner longitudinal (Z) brain position and scanner table (T) position were removed from the whole sample respectively.

### Data analysis

We computed stratified bins by age. The age bins were determined as follows, based on the age at the time of scan (date of the imaging visit minus date of birth): 44-49y, 44≤age<50y; 50-54y, 50≤age<55y; 55-5 y, 55≤age<60y; 60-64y, 60≤age<65y; 65-69y, 65≤age<70y; 70-74y, 70≤age<75y; 75-82y, 75≤age<82y.

#### Validity check

As an initial validation of our data analysis framework, we investigated three brain measures with well-established effects. We used no covariates here because the goal was to evaluate the quality of the raw imaging metrics processed through the UKBB imaging pipeline (Alfaro-Almagro et al., 2018). eTIV, mean cortical thickness and bilateral hippocampal volume are plotted for each age bin split by sex. eTIV, which is a proxy for head size, is expected to be flat across different age bins (Davis and Wright, 1977; Edland et al., 2002; Buckner et al., 2004) but differ by sex. Men show a 12-14% larger head size than women (Blatter et al., 1995; Buckner et al., 2004). Mean cortical thickness is expected to have minimal or no difference between men and women (Im et al., 2008; Holmes et al., 2015), but to decrease with age (Salat et al., 2004). Hippocampal volume is expected to show both an age and a sex effect (Raz et al., 1997; Walhovd et al., 2011). Given historical biases in age-dependent analyses due to the challenge of atrophy (e.g., Pengas et al., 2009), these initial analyses served as a validity check – a form of positive control analysis – that establishes measures behave as they should.

#### Exploration of established AD neurodegenerative biomarkers

To explore the effect of *APOE4* carrier status on well-established AD neurodegenerative biomarkers, we first examined hippocampal volume derived from the T_1_ morphometry and white matter within the CgH derived from dMRI (specifically measured by ICVF). Participants were split by *APOE4* carrier status coding zero, one or two copies of *APOE4* in each age bin. Following prior convention (Elliott et al., 2018), covariates including sex, height, weight at the time of scan, X/Y/Z/T position of head and the radio-frequency receive coil in the scanner, UKBB imaging acquisition center, mean resting-state and task-based functional MRI head motion, inverted SNR in T_1_, inverted CNR in T_1_, FLAIR used or not in the Freesurfer pipeline, eTIV (head size), genotyping chip and the top 40 principal components of the genetic data were regressed. Then, the adjusted bilateral hippocampal volume was plotted by the *APOE4* carrier status. Using the same approach, adjusted ICVF in bilateral CgH was plotted.

To further explore signal properties of the measures, the probability of being an extreme value (in the bottom 5% of the values) was examined after adjusting by age and age^2^. This final analysis allowed exploration of whether carriers of one or two copies of *APOE4* demonstrate an increased probability of having an extreme measured value.

#### Data-driven exploration of novel biomarkers

To identify in a fully data-driven manner candidate AD neurodegenerative biomarkers, 689 IDPs were examined which reflect a diverse array of MRI measures, including 55 T_1_ subcortical volumetric measures from Freesurfer ASEG (Fischl et al., 2002), 202 cortical measures of regional thickness, volume and surface area (Desikan et al., 2006), and 432 dMRI skeleton measurements (48 white matter tract labels x 9 types of dMRI measures for each label) in the JHU ICBM-DTI-81space white matter labels atlas (Mori et al., 2005 and Mori et al., 2007). Based on the signal properties of the two well-established biomarkers which revealed strong effects of *E4/E4* carrier status in the two older age bins, the effect size of carrier status was estimated for each IDP in age bins 65-74y using Cohen’s *d*. Age and age^2^ were regressed from each imaging measure in the exploration of AD effects. A parallel analysis was conducted to estimate effects of aging. The aging effect size was estimated for each imaging measure as the of Pearson’s product moment correlations with age in all the subjects with no copies of *APOE4*.

#### Projections of estimates on cortical surface and dMRI skeleton

To further appreciate the spatial topography of the AD effects and aging effects, visualization of cortical volume data and dMRI measures (using ICVF) was carried out using *wb_view* from the HCP workbench (Marcus et al., 2011). Effect sizes of cortical regions were mapped to a common fsLR32k Desikan-Killiany atlas (Desikan et al., 2006). Effect sizes of dMRI tracts were mapped to a common volumetric JHU ICBM-DTI-81space (Mori et al., 2005 and Mori et al., 2007). A red-white-blue color scale was used to adequately observe the contrast.

## Data Availability

All data analyzed in this study were provided by UK Biobank under application reference 32568. A description of the procedure to access the UK Biobank data is available at: http://www.ukbiobank.ac.uk/using-the-resource/.

## Acknowledgements

This research has been conducted using the UK Biobank resource under applications Number 32568. This work was supported by the Simons Foundation grant 811255, NIH grants R01MH124004, P50MH106435, R00AG054573, and Shared Instrumentation Grant S10OD020039.

## Code Availability

Code used for these analyses will be made available upon publication at the following url: https://github.com/JingnanDu3/Du2021_UKBB_AD

## Author Contributions

J.D. and R.L.B. conceived and designed the study, analyzed the data, and wrote the manuscript. T.G., Z.L., K.M.A, J.F. and L.C.H. also contributed to the data preprocessing, analytic approach or manuscript preparation. All authors contributed to results interpretation and approved the manuscript.

