## Supplemental Tables 1 and 2 for "Exploration of Alzheimer’s Disease MRI Biomarkers Using *APOE4* Carrier Status in the UK Biobank": DuBuckner_SuppTable_20210908.docx

1

Department

of

Psychology,

Center

for

Brain

Science,

Harvard

University,

Cambridge,

MA

02138,

USA

2

The

Computational,

Cognitive

and

Clinical

Neuroimaging

Laboratory,

Hammersmith

Hospital

Campus,

Imperial

College

London,

London

W12

0NN,

UK

3

Athinoula

A.

Martinos

Center

for

Biomedical

Imaging,

Massachusetts

General

Hospital,

Charlestown,

MA

02129,

USA

4

Department

of

Psychiatry,

Massachusetts

General

Hospital,

Charlestown,

MA

02129,

USA

1

Department

of

Psychology,

Center

for

Brain

Science,

Harvard

University,

Cambridge,

MA

02138,

USA

2

The

Computational,

Cognitive

and

Clinical

Neuroimaging

Laboratory,

Hammersmith

Hospital

Campus,

Imperial

College

London,

London

W12

0NN,

UK

3

Athinoula

A.

Martinos

Center

for

Biomedical

Imaging,

Massachusetts

General

Hospital,

Charlestown,

MA

02129,

USA

4

Department

of

Psychiatry,

Massachusetts

General

Hospital,

Charlestown,

MA

02129,

USA

1

Department

of

Psychology,

Center

for

Brain

Science,

Harvard

University,

Cambridge,

MA

02138,

USA

2

The

Computational,

Cognitive

and

Clinical

Neuroimaging

Laboratory,

Hammersmith

Hospital

Campus,

Imperial

College

London,

London

W12

0NN,

UK

3

Athinoula

A.

Martinos

Center

for

Biomedical

Imaging,

Massachusetts

General

Hospital,

Charlestown,

MA

02129,

USA

4

Department

of

Psychiatry,

Massachusetts

General

Hospital,

Charlestown,

MA

02129,

USA

Department

of

Psychology,

Center

for

Brain

Science,

Harvard

University,

Cambridge,

MA

02138,

USA

2

The

Computational,

Cognitive

and

Clinical

Neuroimaging

Laboratory,

Hammersmith

Hospital

Campus,

Imperial

College

London,

London

W12

0NN,

UK

3

Athinoula

A.

Martinos

Center

for

Biomedical

Imaging,

Massachusetts

General

Hospital,

Charlestown,

MA

02129,

USA

4

Department

of

Psychiatry,

Massachusetts

General

Hospital,

Charlestown,

MA

02129,

USA

^1^Department of Psychology, Center for Brain Science, Harvard University, Cambridge, MA 02138, USA; ^2^Department of Psychiatry, Massachusetts General Hospital, Boston, MA 02114, USA; ^3^Psychiatric and Neurodevelopmental Genetics Unit, Center for Genomic Medicine, Massachusetts General Hospital, Boston, MA, 02114, USA; ^4^Institute of Science and Technology for Brain-Inspired Intelligence, Fudan University, Shanghai, China; ^5^Key Laboratory of Computational Neuroscience and Brain-Inspired Intelligence (Fudan University), Ministry of Education, China; ^6^Center for Precision Psychiatry, Massachusetts General Hospital, Boston, MA, 02114, USA; ^7^Stanley Center for Psychiatric Research, Broad Institute of MIT and Harvard, Cambridge, MA, 02138, USA; ^8^Athinoula A. Martinos Center for Biomedical Imaging, Massachusetts General Hospital, Charlestown, MA 02129, USA

**TABLE S1**. The complete list of T1 imaging measures and their estimated *APOE4* effect sizes and aging effect sizes.

| Data Field | T1 structural measures | AD effect sizes | Aging effect sizes |
| --- | --- | --- | --- |
| 26523 | Volume of 3rd-Ventricle (whole brain) | 0.032 | 0.355 |
| 26524 | Volume of 4th-Ventricle (whole brain) | 0.059 | 0.127 |
| 26525 | Volume of 5th-Ventricle (whole brain) | -0.069 | -0.020 |
| 26564 | Volume of Accumbens-area (left hemisphere) | -0.195 | -0.344 |
| 26595 | Volume of Accumbens-area (right hemisphere) | -0.108 | -0.267 |
| 26563 | Volume of Amygdala (left hemisphere) | -0.184 | -0.281 |
| 26594 | Volume of Amygdala (right hemisphere) | -0.222 | -0.217 |
| 26526 | Volume of Brain-Stem (whole brain) | -0.004 | -0.084 |
| 26514 | Volume of BrainSeg (whole brain) | 0.016 | -0.340 |
| 26515 | Volume of BrainSegNotVent (whole brain) | -0.001 | -0.400 |
| 26516 | Volume of BrainSegNotVentSurf (whole brain) | 0.010 | -0.406 |
| 26535 | Volume of CC-Anterior (whole brain) | -0.051 | -0.134 |
| 26533 | Volume of CC-Central (whole brain) | -0.004 | -0.231 |
| 26534 | Volume of CC-Mid-Anterior (whole brain) | -0.053 | -0.253 |
| 26532 | Volume of CC-Mid-Posterior (whole brain) | -0.022 | -0.188 |
| 26531 | Volume of CC-Posterior (whole brain) | -0.044 | -0.008 |
| 26527 | Volume of CSF (whole brain) | -0.005 | 0.191 |
| 26559 | Volume of Caudate (left hemisphere) | -0.067 | 0.000 |
| 26590 | Volume of Caudate (right hemisphere) | -0.077 | 0.026 |
| 26557 | Volume of Cerebellum-Cortex (left hemisphere) | -0.021 | -0.129 |
| 26588 | Volume of Cerebellum-Cortex (right hemisphere) | -0.038 | -0.138 |
| 26556 | Volume of Cerebellum-White-Matter (left hemisphere) | -0.018 | -0.238 |
| 26587 | Volume of Cerebellum-White-Matter (right hemisphere) | -0.018 | -0.208 |
| 26553 | Volume of CerebralWhiteMatter (left hemisphere) | 0.043 | -0.298 |
| 26584 | Volume of CerebralWhiteMatter (right hemisphere) | 0.049 | -0.293 |
| 26552 | Volume of Cortex (left hemisphere) | 0.011 | -0.292 |
| 26583 | Volume of Cortex (right hemisphere) | -0.022 | -0.281 |
| 26562 | Volume of Hippocampus (left hemisphere) | -0.254 | -0.279 |
| 26593 | Volume of Hippocampus (right hemisphere) | -0.254 | -0.271 |
| 26555 | Volume of Inf-Lat-Vent (left hemisphere) | 0.072 | 0.300 |
| 26586 | Volume of Inf-Lat-Vent (right hemisphere) | 0.110 | 0.268 |
| 26554 | Volume of Lateral-Ventricle (left hemisphere) | 0.070 | 0.311 |
| 26585 | Volume of Lateral-Ventricle (right hemisphere) | 0.019 | 0.320 |
| 26530 | Volume of Optic-Chiasm (whole brain) | 0.039 | 0.180 |
| 26561 | Volume of Pallidum (left hemisphere) | -0.073 | -0.134 |
| 26592 | Volume of Pallidum (right hemisphere) | -0.056 | -0.117 |
| 26560 | Volume of Putamen (left hemisphere) | -0.081 | -0.206 |
| 26591 | Volume of Putamen (right hemisphere) | -0.087 | -0.201 |
| 26517 | Volume of SubCortGray (whole brain) | -0.166 | -0.320 |
| 26519 | Volume of SupraTentorial (whole brain) | 0.039 | -0.329 |
| 26520 | Volume of SupraTentorialNotVent (whole brain) | 0.019 | -0.390 |
| 26558 | Volume of Thalamus-Proper (left hemisphere) | -0.053 | -0.337 |
| 26589 | Volume of Thalamus-Proper (right hemisphere) | -0.095 | -0.311 |
| 26518 | Volume of TotalGray (whole brain) | -0.028 | -0.342 |
| 26565 | Volume of VentralDC (left hemisphere) | -0.058 | -0.275 |
| 26596 | Volume of VentralDC (right hemisphere) | -0.034 | -0.288 |
| 26522 | Volume of VentricleChoroid (whole brain) | 0.051 | 0.334 |
| 26528 | Volume of WM-hypointensities (whole brain) | 0.217 | 0.268 |
| 26567 | Volume of choroid-plexus (left hemisphere) | 0.075 | 0.337 |
| 26598 | Volume of choroid-plexus (right hemisphere) | 0.067 | 0.351 |
| 26529 | Volume of non-WM-hypointensities (whole brain) | 0.062 | 0.015 |
| 26566 | Volume of vessel (left hemisphere) | -0.034 | 0.044 |
| 26597 | Volume of vessel (right hemisphere) | 0.004 | 0.069 |
| 26536 | Volume-ratio of BrainSegVol-to-eTIV (whole brain) | 0.001 | -0.332 |
| 26537 | Volume-ratio of MaskVol-to-eTIV (whole brain) | -0.071 | -0.176 |
| 26755 | Mean thickness of GlobalMeanMean thickness (left hemisphere) | -0.020 | -0.264 |
| 26856 | Mean thickness of GlobalMeanMean thickness (right hemisphere) | -0.061 | -0.255 |
| 26756 | Mean thickness of bankssts (left hemisphere) | 0.047 | -0.127 |
| 26857 | Mean thickness of bankssts (right hemisphere) | 0.022 | -0.139 |
| 26757 | Mean thickness of caudalanteriorcingulate (left hemisphere) | -0.023 | -0.037 |
| 26858 | Mean thickness of caudalanteriorcingulate (right hemisphere) | -0.032 | -0.028 |
| 26758 | Mean thickness of caudalmiddlefrontal (left hemisphere) | -0.020 | -0.224 |
| 26859 | Mean thickness of caudalmiddlefrontal (right hemisphere) | -0.122 | -0.219 |
| 26759 | Mean thickness of cuneus (left hemisphere) | 0.048 | -0.058 |
| 26860 | Mean thickness of cuneus (right hemisphere) | 0.024 | -0.008 |
| 26760 | Mean thickness of entorhinal (left hemisphere) | -0.019 | -0.106 |
| 26861 | Mean thickness of entorhinal (right hemisphere) | -0.154 | -0.099 |
| 26786 | Mean thickness of frontalpole (left hemisphere) | 0.016 | -0.117 |
| 26887 | Mean thickness of frontalpole (right hemisphere) | 0.027 | -0.107 |
| 26761 | Mean thickness of fusiform (left hemisphere) | 0.041 | -0.164 |
| 26862 | Mean thickness of fusiform (right hemisphere) | -0.021 | -0.177 |
| 26762 | Mean thickness of inferiorparietal (left hemisphere) | -0.066 | -0.255 |
| 26863 | Mean thickness of inferiorparietal (right hemisphere) | -0.060 | -0.270 |
| 26763 | Mean thickness of inferiortemporal (left hemisphere) | 0.000 | -0.107 |
| 26864 | Mean thickness of inferiortemporal (right hemisphere) | -0.059 | -0.136 |
| 26788 | Mean thickness of insula (left hemisphere) | 0.032 | -0.092 |
| 26889 | Mean thickness of insula (right hemisphere) | -0.020 | -0.097 |
| 26764 | Mean thickness of isthmuscingulate (left hemisphere) | -0.029 | -0.136 |
| 26865 | Mean thickness of isthmuscingulate (right hemisphere) | -0.043 | -0.117 |
| 26765 | Mean thickness of lateraloccipital (left hemisphere) | 0.004 | -0.127 |
| 26866 | Mean thickness of lateraloccipital (right hemisphere) | -0.078 | -0.121 |
| 26766 | Mean thickness of lateralorbitofrontal (left hemisphere) | -0.019 | -0.103 |
| 26867 | Mean thickness of lateralorbitofrontal (right hemisphere) | 0.038 | -0.079 |
| 26767 | Mean thickness of lingual (left hemisphere) | -0.070 | -0.103 |
| 26868 | Mean thickness of lingual (right hemisphere) | -0.035 | -0.056 |
| 26768 | Mean thickness of medialorbitofrontal (left hemisphere) | -0.140 | -0.113 |
| 26869 | Mean thickness of medialorbitofrontal (right hemisphere) | -0.147 | -0.082 |
| 26769 | Mean thickness of middletemporal (left hemisphere) | -0.111 | -0.156 |
| 26870 | Mean thickness of middletemporal (right hemisphere) | -0.135 | -0.169 |
| 26771 | Mean thickness of paracentral (left hemisphere) | -0.062 | -0.211 |
| 26872 | Mean thickness of paracentral (right hemisphere) | -0.084 | -0.198 |
| 26770 | Mean thickness of parahippocampal (left hemisphere) | -0.002 | -0.080 |
| 26871 | Mean thickness of parahippocampal (right hemisphere) | -0.065 | -0.080 |
| 26772 | Mean thickness of parsopercularis (left hemisphere) | 0.081 | -0.223 |
| 26873 | Mean thickness of parsopercularis (right hemisphere) | -0.009 | -0.203 |
| 26773 | Mean thickness of parsorbitalis (left hemisphere) | 0.021 | -0.158 |
| 26874 | Mean thickness of parsorbitalis (right hemisphere) | -0.024 | -0.161 |
| 26774 | Mean thickness of parstriangularis (left hemisphere) | 0.013 | -0.235 |
| 26875 | Mean thickness of parstriangularis (right hemisphere) | 0.025 | -0.234 |
| 26775 | Mean thickness of pericalcarine (left hemisphere) | 0.055 | -0.038 |
| 26876 | Mean thickness of pericalcarine (right hemisphere) | -0.085 | 0.019 |
| 26776 | Mean thickness of postcentral (left hemisphere) | 0.071 | -0.209 |
| 26877 | Mean thickness of postcentral (right hemisphere) | 0.033 | -0.200 |
| 26777 | Mean thickness of posteriorcingulate (left hemisphere) | 0.066 | -0.098 |
| 26878 | Mean thickness of posteriorcingulate (right hemisphere) | -0.030 | -0.102 |
| 26778 | Mean thickness of precentral (left hemisphere) | -0.019 | -0.242 |
| 26879 | Mean thickness of precentral (right hemisphere) | -0.069 | -0.241 |
| 26779 | Mean thickness of precuneus (left hemisphere) | 0.000 | -0.257 |
| 26880 | Mean thickness of precuneus (right hemisphere) | -0.036 | -0.241 |
| 26780 | Mean thickness of rostralanteriorcingulate (left hemisphere) | -0.009 | -0.106 |
| 26881 | Mean thickness of rostralanteriorcingulate (right hemisphere) | 0.009 | -0.019 |
| 26781 | Mean thickness of rostralmiddlefrontal (left hemisphere) | -0.039 | -0.271 |
| 26882 | Mean thickness of rostralmiddlefrontal (right hemisphere) | -0.052 | -0.249 |
| 26782 | Mean thickness of superiorfrontal (left hemisphere) | -0.085 | -0.310 |
| 26883 | Mean thickness of superiorfrontal (right hemisphere) | -0.085 | -0.298 |
| 26783 | Mean thickness of superiorparietal (left hemisphere) | 0.088 | -0.234 |
| 26884 | Mean thickness of superiorparietal (right hemisphere) | 0.067 | -0.239 |
| 26784 | Mean thickness of superiortemporal (left hemisphere) | -0.023 | -0.242 |
| 26885 | Mean thickness of superiortemporal (right hemisphere) | -0.050 | -0.266 |
| 26785 | Mean thickness of supramarginal (left hemisphere) | -0.067 | -0.261 |
| 26886 | Mean thickness of supramarginal (right hemisphere) | -0.045 | -0.251 |
| 26787 | Mean thickness of transversetemporal (left hemisphere) | 0.067 | -0.008 |
| 26888 | Mean thickness of transversetemporal (right hemisphere) | 0.027 | -0.022 |
| 26721 | Area of TotalSurface (left hemisphere) | 0.034 | -0.111 |
| 26822 | Area of TotalSurface (right hemisphere) | 0.038 | -0.100 |
| 26722 | Area of bankssts (left hemisphere) | -0.036 | -0.039 |
| 26823 | Area of bankssts (right hemisphere) | -0.008 | -0.052 |
| 26723 | Area of caudalanteriorcingulate (left hemisphere) | -0.105 | -0.023 |
| 26824 | Area of caudalanteriorcingulate (right hemisphere) | 0.014 | -0.025 |
| 26724 | Area of caudalmiddlefrontal (left hemisphere) | -0.027 | -0.025 |
| 26825 | Area of caudalmiddlefrontal (right hemisphere) | -0.075 | -0.010 |
| 26725 | Area of cuneus (left hemisphere) | 0.010 | -0.081 |
| 26826 | Area of cuneus (right hemisphere) | 0.069 | -0.078 |
| 26726 | Area of entorhinal (left hemisphere) | 0.007 | 0.003 |
| 26827 | Area of entorhinal (right hemisphere) | 0.004 | 0.012 |
| 26752 | Area of frontalpole (left hemisphere) | -0.109 | -0.091 |
| 26853 | Area of frontalpole (right hemisphere) | -0.099 | -0.052 |
| 26727 | Area of fusiform (left hemisphere) | -0.022 | -0.101 |
| 26828 | Area of fusiform (right hemisphere) | 0.036 | -0.079 |
| 26728 | Area of inferiorparietal (left hemisphere) | 0.014 | -0.032 |
| 26829 | Area of inferiorparietal (right hemisphere) | 0.023 | -0.043 |
| 26729 | Area of inferiortemporal (left hemisphere) | -0.068 | -0.108 |
| 26830 | Area of inferiortemporal (right hemisphere) | 0.031 | -0.085 |
| 26754 | Area of insula (left hemisphere) | 0.017 | 0.023 |
| 26855 | Area of insula (right hemisphere) | -0.031 | 0.015 |
| 26730 | Area of isthmuscingulate (left hemisphere) | -0.076 | 0.030 |
| 26831 | Area of isthmuscingulate (right hemisphere) | -0.042 | 0.021 |
| 26731 | Area of lateraloccipital (left hemisphere) | 0.056 | -0.096 |
| 26832 | Area of lateraloccipital (right hemisphere) | 0.033 | -0.086 |
| 26732 | Area of lateralorbitofrontal (left hemisphere) | 0.021 | -0.115 |
| 26833 | Area of lateralorbitofrontal (right hemisphere) | -0.015 | -0.084 |
| 26733 | Area of lingual (left hemisphere) | 0.042 | -0.072 |
| 26834 | Area of lingual (right hemisphere) | 0.037 | -0.060 |
| 26734 | Area of medialorbitofrontal (left hemisphere) | -0.069 | -0.031 |
| 26835 | Area of medialorbitofrontal (right hemisphere) | -0.013 | -0.050 |
| 26735 | Area of middletemporal (left hemisphere) | -0.055 | -0.097 |
| 26836 | Area of middletemporal (right hemisphere) | -0.106 | -0.099 |
| 26737 | Area of paracentral (left hemisphere) | 0.113 | 0.022 |
| 26838 | Area of paracentral (right hemisphere) | 0.173 | 0.015 |
| 26736 | Area of parahippocampal (left hemisphere) | 0.020 | -0.112 |
| 26837 | Area of parahippocampal (right hemisphere) | 0.008 | -0.089 |
| 26738 | Area of parsopercularis (left hemisphere) | -0.129 | -0.058 |
| 26839 | Area of parsopercularis (right hemisphere) | 0.011 | -0.043 |
| 26739 | Area of parsorbitalis (left hemisphere) | 0.044 | -0.098 |
| 26840 | Area of parsorbitalis (right hemisphere) | 0.035 | -0.103 |
| 26740 | Area of parstriangularis (left hemisphere) | -0.014 | -0.058 |
| 26841 | Area of parstriangularis (right hemisphere) | 0.046 | -0.064 |
| 26741 | Area of pericalcarine (left hemisphere) | 0.012 | -0.037 |
| 26842 | Area of pericalcarine (right hemisphere) | 0.032 | -0.043 |
| 26742 | Area of postcentral (left hemisphere) | 0.029 | -0.010 |
| 26843 | Area of postcentral (right hemisphere) | 0.040 | 0.015 |
| 26743 | Area of posteriorcingulate (left hemisphere) | -0.062 | -0.046 |
| 26844 | Area of posteriorcingulate (right hemisphere) | 0.022 | -0.059 |
| 26744 | Area of precentral (left hemisphere) | 0.103 | 0.007 |
| 26845 | Area of precentral (right hemisphere) | 0.077 | 0.032 |
| 26745 | Area of precuneus (left hemisphere) | 0.002 | -0.060 |
| 26846 | Area of precuneus (right hemisphere) | -0.061 | -0.056 |
| 26746 | Area of rostralanteriorcingulate (left hemisphere) | 0.021 | 0.004 |
| 26847 | Area of rostralanteriorcingulate (right hemisphere) | 0.032 | -0.024 |
| 26747 | Area of rostralmiddlefrontal (left hemisphere) | 0.067 | -0.073 |
| 26848 | Area of rostralmiddlefrontal (right hemisphere) | 0.014 | -0.081 |
| 26748 | Area of superiorfrontal (left hemisphere) | 0.089 | -0.042 |
| 26849 | Area of superiorfrontal (right hemisphere) | 0.049 | -0.044 |
| 26749 | Area of superiorparietal (left hemisphere) | 0.088 | -0.047 |
| 26850 | Area of superiorparietal (right hemisphere) | 0.036 | -0.048 |
| 26750 | Area of superiortemporal (left hemisphere) | -0.005 | -0.034 |
| 26851 | Area of superiortemporal (right hemisphere) | -0.006 | -0.024 |
| 26751 | Area of supramarginal (left hemisphere) | -0.018 | -0.012 |
| 26852 | Area of supramarginal (right hemisphere) | 0.024 | 0.011 |
| 26753 | Area of transversetemporal (left hemisphere) | 0.062 | -0.059 |
| 26854 | Area of transversetemporal (right hemisphere) | 0.040 | -0.055 |
| 26789 | Volume of bankssts (left hemisphere) | -0.016 | -0.093 |
| 26890 | Volume of bankssts (right hemisphere) | 0.026 | -0.108 |
| 26790 | Volume of caudalanteriorcingulate (left hemisphere) | -0.095 | -0.061 |
| 26891 | Volume of caudalanteriorcingulate (right hemisphere) | 0.048 | -0.058 |
| 26791 | Volume of caudalmiddlefrontal (left hemisphere) | -0.054 | -0.132 |
| 26892 | Volume of caudalmiddlefrontal (right hemisphere) | -0.155 | -0.112 |
| 26792 | Volume of cuneus (left hemisphere) | 0.034 | -0.084 |
| 26893 | Volume of cuneus (right hemisphere) | 0.074 | -0.044 |
| 26793 | Volume of entorhinal (left hemisphere) | -0.027 | -0.027 |
| 26894 | Volume of entorhinal (right hemisphere) | -0.071 | -0.014 |
| 26819 | Volume of frontalpole (left hemisphere) | -0.045 | -0.109 |
| 26920 | Volume of frontalpole (right hemisphere) | -0.002 | -0.087 |
| 26794 | Volume of fusiform (left hemisphere) | 0.020 | -0.160 |
| 26895 | Volume of fusiform (right hemisphere) | -0.004 | -0.164 |
| 26795 | Volume of inferiorparietal (left hemisphere) | -0.006 | -0.135 |
| 26896 | Volume of inferiorparietal (right hemisphere) | -0.007 | -0.176 |
| 26796 | Volume of inferiortemporal (left hemisphere) | 0.027 | -0.123 |
| 26897 | Volume of inferiortemporal (right hemisphere) | 0.076 | -0.131 |
| 26821 | Volume of insula (left hemisphere) | 0.091 | 0.000 |
| 26922 | Volume of insula (right hemisphere) | -0.010 | -0.016 |
| 26797 | Volume of isthmuscingulate (left hemisphere) | -0.118 | -0.048 |
| 26898 | Volume of isthmuscingulate (right hemisphere) | -0.066 | -0.045 |
| 26798 | Volume of lateraloccipital (left hemisphere) | 0.045 | -0.150 |
| 26899 | Volume of lateraloccipital (right hemisphere) | -0.009 | -0.143 |
| 26799 | Volume of lateralorbitofrontal (left hemisphere) | 0.034 | -0.173 |
| 26900 | Volume of lateralorbitofrontal (right hemisphere) | 0.044 | -0.150 |
| 26800 | Volume of lingual (left hemisphere) | -0.006 | -0.106 |
| 26901 | Volume of lingual (right hemisphere) | 0.024 | -0.070 |
| 26801 | Volume of medialorbitofrontal (left hemisphere) | -0.132 | -0.126 |
| 26902 | Volume of medialorbitofrontal (right hemisphere) | -0.061 | -0.140 |
| 26802 | Volume of middletemporal (left hemisphere) | -0.038 | -0.172 |
| 26903 | Volume of middletemporal (right hemisphere) | -0.120 | -0.181 |
| 26804 | Volume of paracentral (left hemisphere) | 0.019 | -0.146 |
| 26905 | Volume of paracentral (right hemisphere) | 0.064 | -0.130 |
| 26803 | Volume of parahippocampal (left hemisphere) | 0.015 | -0.124 |
| 26904 | Volume of parahippocampal (right hemisphere) | -0.045 | -0.107 |
| 26805 | Volume of parsopercularis (left hemisphere) | -0.107 | -0.152 |
| 26906 | Volume of parsopercularis (right hemisphere) | -0.024 | -0.139 |
| 26806 | Volume of parsorbitalis (left hemisphere) | 0.068 | -0.172 |
| 26907 | Volume of parsorbitalis (right hemisphere) | 0.059 | -0.186 |
| 26807 | Volume of parstriangularis (left hemisphere) | -0.017 | -0.158 |
| 26908 | Volume of parstriangularis (right hemisphere) | 0.044 | -0.161 |
| 26808 | Volume of pericalcarine (left hemisphere) | 0.013 | -0.020 |
| 26909 | Volume of pericalcarine (right hemisphere) | -0.030 | 0.011 |
| 26809 | Volume of postcentral (left hemisphere) | 0.050 | -0.147 |
| 26910 | Volume of postcentral (right hemisphere) | 0.036 | -0.131 |
| 26810 | Volume of posteriorcingulate (left hemisphere) | -0.023 | -0.074 |
| 26911 | Volume of posteriorcingulate (right hemisphere) | 0.021 | -0.103 |
| 26811 | Volume of precentral (left hemisphere) | 0.029 | -0.192 |
| 26912 | Volume of precentral (right hemisphere) | -0.044 | -0.174 |
| 26812 | Volume of precuneus (left hemisphere) | -0.010 | -0.209 |
| 26913 | Volume of precuneus (right hemisphere) | -0.104 | -0.186 |
| 26813 | Volume of rostralanteriorcingulate (left hemisphere) | 0.053 | -0.056 |
| 26914 | Volume of rostralanteriorcingulate (right hemisphere) | 0.083 | -0.040 |
| 26814 | Volume of rostralmiddlefrontal (left hemisphere) | 0.041 | -0.210 |
| 26915 | Volume of rostralmiddlefrontal (right hemisphere) | -0.008 | -0.196 |
| 26815 | Volume of superiorfrontal (left hemisphere) | 0.034 | -0.222 |
| 26916 | Volume of superiorfrontal (right hemisphere) | 0.007 | -0.210 |
| 26816 | Volume of superiorparietal (left hemisphere) | 0.118 | -0.172 |
| 26917 | Volume of superiorparietal (right hemisphere) | 0.048 | -0.184 |
| 26817 | Volume of superiortemporal (left hemisphere) | -0.002 | -0.168 |
| 26918 | Volume of superiortemporal (right hemisphere) | -0.018 | -0.172 |
| 26818 | Volume of supramarginal (left hemisphere) | -0.051 | -0.116 |
| 26919 | Volume of supramarginal (right hemisphere) | 0.002 | -0.106 |
| 26820 | Volume of transversetemporal (left hemisphere) | 0.108 | -0.036 |
| 26921 | Volume of transversetemporal (right hemisphere) | 0.044 | -0.037 |

**TABLE S2**. The complete list of DTI measures and their estimated *APOE4* effect sizes and aging effect sizes.

| Data Field | DTI measures | AD effect sizes | Aging effect sizes |
| --- | --- | --- | --- |
| 25079 | Mean FA in anterior corona radiata on FA skeleton (left) | -0.165 | -0.338 |
| 25078 | Mean FA in anterior corona radiata on FA skeleton (right) | -0.119 | -0.291 |
| 25073 | Mean FA in anterior limb of internal capsule on FA skeleton (left) | -0.037 | -0.182 |
| 25072 | Mean FA in anterior limb of internal capsule on FA skeleton (right) | -0.052 | -0.179 |
| 25059 | Mean FA in body of corpus callosum on FA skeleton | -0.145 | -0.233 |
| 25071 | Mean FA in cerebral peduncle on FA skeleton (left) | -0.172 | -0.229 |
| 25070 | Mean FA in cerebral peduncle on FA skeleton (right) | -0.200 | -0.183 |
| 25091 | Mean FA in cingulum cingulate gyrus on FA skeleton (left) | -0.083 | -0.214 |
| 25090 | Mean FA in cingulum cingulate gyrus on FA skeleton (right) | -0.037 | -0.205 |
| 25093 | Mean FA in cingulum hippocampus on FA skeleton (left) | -0.245 | -0.114 |
| 25092 | Mean FA in cingulum hippocampus on FA skeleton (right) | -0.276 | -0.089 |
| 25063 | Mean FA in corticospinal tract on FA skeleton (left) | -0.138 | -0.127 |
| 25062 | Mean FA in corticospinal tract on FA skeleton (right) | -0.155 | -0.101 |
| 25089 | Mean FA in external capsule on FA skeleton (left) | -0.064 | -0.267 |
| 25088 | Mean FA in external capsule on FA skeleton (right) | -0.119 | -0.286 |
| 25095 | Mean FA in fornix cres+stria terminalis on FA skeleton (left) | -0.042 | -0.359 |
| 25094 | Mean FA in fornix cres+stria terminalis on FA skeleton (right) | -0.094 | -0.364 |
| 25061 | Mean FA in fornix on FA skeleton | -0.063 | -0.392 |
| 25058 | Mean FA in genu of corpus callosum on FA skeleton | -0.105 | -0.275 |
| 25067 | Mean FA in inferior cerebellar peduncle on FA skeleton (left) | -0.106 | -0.156 |
| 25066 | Mean FA in inferior cerebellar peduncle on FA skeleton (right) | -0.109 | -0.180 |
| 25065 | Mean FA in medial lemniscus on FA skeleton (left) | -0.116 | -0.084 |
| 25064 | Mean FA in medial lemniscus on FA skeleton (right) | -0.080 | -0.089 |
| 25056 | Mean FA in middle cerebellar peduncle on FA skeleton | -0.272 | -0.149 |
| 25057 | Mean FA in pontine crossing tract on FA skeleton | -0.083 | -0.082 |
| 25083 | Mean FA in posterior corona radiata on FA skeleton (left) | -0.161 | -0.162 |
| 25082 | Mean FA in posterior corona radiata on FA skeleton (right) | -0.170 | -0.114 |
| 25075 | Mean FA in posterior limb of internal capsule on FA skeleton (left) | -0.024 | -0.052 |
| 25074 | Mean FA in posterior limb of internal capsule on FA skeleton (right) | -0.060 | -0.058 |
| 25085 | Mean FA in posterior thalamic radiation on FA skeleton (left) | -0.288 | -0.256 |
| 25084 | Mean FA in posterior thalamic radiation on FA skeleton (right) | -0.352 | -0.263 |
| 25077 | Mean FA in retrolenticular part of internal capsule on FA skeleton (left) | -0.071 | -0.112 |
| 25076 | Mean FA in retrolenticular part of internal capsule on FA skeleton (right) | -0.162 | -0.082 |
| 25087 | Mean FA in sagittal stratum on FA skeleton (left) | -0.263 | -0.159 |
| 25086 | Mean FA in sagittal stratum on FA skeleton (right) | -0.303 | -0.169 |
| 25060 | Mean FA in splenium of corpus callosum on FA skeleton | -0.174 | -0.088 |
| 25069 | Mean FA in superior cerebellar peduncle on FA skeleton (left) | 0.005 | 0.000 |
| 25068 | Mean FA in superior cerebellar peduncle on FA skeleton (right) | -0.021 | 0.016 |
| 25081 | Mean FA in superior corona radiata on FA skeleton (left) | -0.104 | -0.198 |
| 25080 | Mean FA in superior corona radiata on FA skeleton (right) | -0.132 | -0.180 |
| 25099 | Mean FA in superior fronto-occipital fasciculus on FA skeleton (left) | -0.078 | -0.271 |
| 25098 | Mean FA in superior fronto-occipital fasciculus on FA skeleton (right) | -0.147 | -0.245 |
| 25097 | Mean FA in superior longitudinal fasciculus on FA skeleton (left) | -0.128 | -0.178 |
| 25096 | Mean FA in superior longitudinal fasciculus on FA skeleton (right) | -0.158 | -0.169 |
| 25103 | Mean FA in tapetum on FA skeleton (left) | 0.000 | -0.126 |
| 25102 | Mean FA in tapetum on FA skeleton (right) | -0.160 | -0.193 |
| 25101 | Mean FA in uncinate fasciculus on FA skeleton (left) | -0.062 | -0.116 |
| 25100 | Mean FA in uncinate fasciculus on FA skeleton (right) | -0.164 | -0.105 |
| 25367 | Mean ICVF in anterior corona radiata on FA skeleton (left) | -0.169 | -0.304 |
| 25366 | Mean ICVF in anterior corona radiata on FA skeleton (right) | -0.179 | -0.308 |
| 25361 | Mean ICVF in anterior limb of internal capsule on FA skeleton (left) | -0.077 | -0.253 |
| 25360 | Mean ICVF in anterior limb of internal capsule on FA skeleton (right) | -0.085 | -0.264 |
| 25347 | Mean ICVF in body of corpus callosum on FA skeleton | -0.119 | -0.065 |
| 25359 | Mean ICVF in cerebral peduncle on FA skeleton (left) | -0.074 | -0.022 |
| 25358 | Mean ICVF in cerebral peduncle on FA skeleton (right) | -0.099 | -0.003 |
| 25379 | Mean ICVF in cingulum cingulate gyrus on FA skeleton (left) | -0.136 | -0.246 |
| 25378 | Mean ICVF in cingulum cingulate gyrus on FA skeleton (right) | -0.090 | -0.241 |
| 25381 | Mean ICVF in cingulum hippocampus on FA skeleton (left) | -0.344 | -0.139 |
| 25380 | Mean ICVF in cingulum hippocampus on FA skeleton (right) | -0.302 | -0.138 |
| 25351 | Mean ICVF in corticospinal tract on FA skeleton (left) | -0.061 | 0.037 |
| 25350 | Mean ICVF in corticospinal tract on FA skeleton (right) | -0.115 | 0.043 |
| 25377 | Mean ICVF in external capsule on FA skeleton (left) | -0.134 | -0.297 |
| 25376 | Mean ICVF in external capsule on FA skeleton (right) | -0.144 | -0.293 |
| 25383 | Mean ICVF in fornix cres+stria terminalis on FA skeleton (left) | -0.022 | -0.155 |
| 25382 | Mean ICVF in fornix cres+stria terminalis on FA skeleton (right) | -0.028 | -0.133 |
| 25349 | Mean ICVF in fornix on FA skeleton | -0.138 | -0.175 |
| 25346 | Mean ICVF in genu of corpus callosum on FA skeleton | -0.095 | -0.208 |
| 25355 | Mean ICVF in inferior cerebellar peduncle on FA skeleton (left) | -0.021 | -0.051 |
| 25354 | Mean ICVF in inferior cerebellar peduncle on FA skeleton (right) | -0.002 | -0.040 |
| 25353 | Mean ICVF in medial lemniscus on FA skeleton (left) | -0.011 | 0.006 |
| 25352 | Mean ICVF in medial lemniscus on FA skeleton (right) | -0.024 | 0.012 |
| 25344 | Mean ICVF in middle cerebellar peduncle on FA skeleton | -0.070 | -0.095 |
| 25345 | Mean ICVF in pontine crossing tract on FA skeleton | 0.002 | -0.005 |
| 25371 | Mean ICVF in posterior corona radiata on FA skeleton (left) | -0.218 | -0.212 |
| 25370 | Mean ICVF in posterior corona radiata on FA skeleton (right) | -0.236 | -0.222 |
| 25363 | Mean ICVF in posterior limb of internal capsule on FA skeleton (left) | -0.059 | -0.175 |
| 25362 | Mean ICVF in posterior limb of internal capsule on FA skeleton (right) | -0.055 | -0.168 |
| 25373 | Mean ICVF in posterior thalamic radiation on FA skeleton (left) | -0.301 | -0.199 |
| 25372 | Mean ICVF in posterior thalamic radiation on FA skeleton (right) | -0.365 | -0.180 |
| 25365 | Mean ICVF in retrolenticular part of internal capsule on FA skeleton (left) | -0.088 | -0.148 |
| 25364 | Mean ICVF in retrolenticular part of internal capsule on FA skeleton (right) | -0.137 | -0.156 |
| 25375 | Mean ICVF in sagittal stratum on FA skeleton (left) | -0.290 | -0.185 |
| 25374 | Mean ICVF in sagittal stratum on FA skeleton (right) | -0.267 | -0.166 |
| 25348 | Mean ICVF in splenium of corpus callosum on FA skeleton | -0.166 | -0.161 |
| 25357 | Mean ICVF in superior cerebellar peduncle on FA skeleton (left) | -0.001 | 0.042 |
| 25356 | Mean ICVF in superior cerebellar peduncle on FA skeleton (right) | 0.014 | 0.071 |
| 25369 | Mean ICVF in superior corona radiata on FA skeleton (left) | -0.138 | -0.262 |
| 25368 | Mean ICVF in superior corona radiata on FA skeleton (right) | -0.155 | -0.246 |
| 25387 | Mean ICVF in superior fronto-occipital fasciculus on FA skeleton (left) | -0.101 | -0.361 |
| 25386 | Mean ICVF in superior fronto-occipital fasciculus on FA skeleton (right) | -0.114 | -0.347 |
| 25385 | Mean ICVF in superior longitudinal fasciculus on FA skeleton (left) | -0.154 | -0.208 |
| 25384 | Mean ICVF in superior longitudinal fasciculus on FA skeleton (right) | -0.169 | -0.181 |
| 25391 | Mean ICVF in tapetum on FA skeleton (left) | -0.167 | -0.191 |
| 25390 | Mean ICVF in tapetum on FA skeleton (right) | -0.210 | -0.216 |
| 25389 | Mean ICVF in uncinate fasciculus on FA skeleton (left) | -0.067 | -0.227 |
| 25388 | Mean ICVF in uncinate fasciculus on FA skeleton (right) | -0.092 | -0.225 |
| 25463 | Mean ISOVF in anterior corona radiata on FA skeleton (left) | 0.138 | 0.159 |
| 25462 | Mean ISOVF in anterior corona radiata on FA skeleton (right) | 0.086 | 0.111 |
| 25457 | Mean ISOVF in anterior limb of internal capsule on FA skeleton (left) | 0.027 | 0.220 |
| 25456 | Mean ISOVF in anterior limb of internal capsule on FA skeleton (right) | -0.015 | 0.207 |
| 25443 | Mean ISOVF in body of corpus callosum on FA skeleton | 0.150 | 0.323 |
| 25455 | Mean ISOVF in cerebral peduncle on FA skeleton (left) | 0.049 | 0.127 |
| 25454 | Mean ISOVF in cerebral peduncle on FA skeleton (right) | 0.079 | 0.128 |
| 25475 | Mean ISOVF in cingulum cingulate gyrus on FA skeleton (left) | 0.141 | -0.046 |
| 25474 | Mean ISOVF in cingulum cingulate gyrus on FA skeleton (right) | 0.150 | -0.055 |
| 25477 | Mean ISOVF in cingulum hippocampus on FA skeleton (left) | -0.066 | 0.152 |
| 25476 | Mean ISOVF in cingulum hippocampus on FA skeleton (right) | 0.014 | 0.092 |
| 25447 | Mean ISOVF in corticospinal tract on FA skeleton (left) | 0.092 | 0.036 |
| 25446 | Mean ISOVF in corticospinal tract on FA skeleton (right) | 0.052 | 0.016 |
| 25473 | Mean ISOVF in external capsule on FA skeleton (left) | 0.043 | 0.208 |
| 25472 | Mean ISOVF in external capsule on FA skeleton (right) | 0.095 | 0.188 |
| 25479 | Mean ISOVF in fornix cres+stria terminalis on FA skeleton (left) | 0.010 | 0.232 |
| 25478 | Mean ISOVF in fornix cres+stria terminalis on FA skeleton (right) | -0.008 | 0.277 |
| 25445 | Mean ISOVF in fornix on FA skeleton | 0.003 | 0.414 |
| 25442 | Mean ISOVF in genu of corpus callosum on FA skeleton | 0.092 | 0.273 |
| 25451 | Mean ISOVF in inferior cerebellar peduncle on FA skeleton (left) | 0.097 | 0.128 |
| 25450 | Mean ISOVF in inferior cerebellar peduncle on FA skeleton (right) | 0.118 | 0.130 |
| 25449 | Mean ISOVF in medial lemniscus on FA skeleton (left) | 0.052 | 0.035 |
| 25448 | Mean ISOVF in medial lemniscus on FA skeleton (right) | 0.058 | 0.033 |
| 25440 | Mean ISOVF in middle cerebellar peduncle on FA skeleton | 0.072 | 0.034 |
| 25441 | Mean ISOVF in pontine crossing tract on FA skeleton | 0.089 | -0.004 |
| 25467 | Mean ISOVF in posterior corona radiata on FA skeleton (left) | 0.146 | 0.193 |
| 25466 | Mean ISOVF in posterior corona radiata on FA skeleton (right) | 0.139 | 0.217 |
| 25459 | Mean ISOVF in posterior limb of internal capsule on FA skeleton (left) | 0.027 | 0.116 |
| 25458 | Mean ISOVF in posterior limb of internal capsule on FA skeleton (right) | 0.057 | 0.147 |
| 25469 | Mean ISOVF in posterior thalamic radiation on FA skeleton (left) | 0.369 | 0.176 |
| 25468 | Mean ISOVF in posterior thalamic radiation on FA skeleton (right) | 0.395 | 0.155 |
| 25461 | Mean ISOVF in retrolenticular part of internal capsule on FA skeleton (left) | 0.016 | 0.114 |
| 25460 | Mean ISOVF in retrolenticular part of internal capsule on FA skeleton (right) | 0.116 | 0.112 |
| 25471 | Mean ISOVF in sagittal stratum on FA skeleton (left) | 0.219 | 0.104 |
| 25470 | Mean ISOVF in sagittal stratum on FA skeleton (right) | 0.277 | 0.111 |
| 25444 | Mean ISOVF in splenium of corpus callosum on FA skeleton | 0.080 | 0.140 |
| 25453 | Mean ISOVF in superior cerebellar peduncle on FA skeleton (left) | -0.012 | 0.167 |
| 25452 | Mean ISOVF in superior cerebellar peduncle on FA skeleton (right) | -0.006 | 0.166 |
| 25465 | Mean ISOVF in superior corona radiata on FA skeleton (left) | 0.214 | 0.246 |
| 25464 | Mean ISOVF in superior corona radiata on FA skeleton (right) | 0.238 | 0.259 |
| 25483 | Mean ISOVF in superior fronto-occipital fasciculus on FA skeleton (left) | 0.000 | 0.194 |
| 25482 | Mean ISOVF in superior fronto-occipital fasciculus on FA skeleton (right) | 0.106 | 0.166 |
| 25481 | Mean ISOVF in superior longitudinal fasciculus on FA skeleton (left) | 0.297 | 0.159 |
| 25480 | Mean ISOVF in superior longitudinal fasciculus on FA skeleton (right) | 0.265 | 0.211 |
| 25487 | Mean ISOVF in tapetum on FA skeleton (left) | -0.009 | 0.157 |
| 25486 | Mean ISOVF in tapetum on FA skeleton (right) | 0.117 | 0.212 |
| 25485 | Mean ISOVF in uncinate fasciculus on FA skeleton (left) | 0.027 | 0.074 |
| 25484 | Mean ISOVF in uncinate fasciculus on FA skeleton (right) | 0.320 | 0.056 |
| 25223 | Mean L1 in anterior corona radiata on FA skeleton (left) | 0.191 | 0.223 |
| 25222 | Mean L1 in anterior corona radiata on FA skeleton (right) | 0.201 | 0.231 |
| 25217 | Mean L1 in anterior limb of internal capsule on FA skeleton (left) | 0.065 | 0.318 |
| 25216 | Mean L1 in anterior limb of internal capsule on FA skeleton (right) | 0.015 | 0.317 |
| 25203 | Mean L1 in body of corpus callosum on FA skeleton | 0.107 | 0.199 |
| 25215 | Mean L1 in cerebral peduncle on FA skeleton (left) | -0.020 | -0.085 |
| 25214 | Mean L1 in cerebral peduncle on FA skeleton (right) | 0.014 | -0.045 |
| 25235 | Mean L1 in cingulum cingulate gyrus on FA skeleton (left) | 0.177 | -0.030 |
| 25234 | Mean L1 in cingulum cingulate gyrus on FA skeleton (right) | 0.163 | -0.031 |
| 25237 | Mean L1 in cingulum hippocampus on FA skeleton (left) | 0.083 | -0.007 |
| 25236 | Mean L1 in cingulum hippocampus on FA skeleton (right) | 0.038 | 0.001 |
| 25207 | Mean L1 in corticospinal tract on FA skeleton (left) | 0.053 | -0.052 |
| 25206 | Mean L1 in corticospinal tract on FA skeleton (right) | 0.023 | -0.061 |
| 25233 | Mean L1 in external capsule on FA skeleton (left) | 0.140 | 0.248 |
| 25232 | Mean L1 in external capsule on FA skeleton (right) | 0.154 | 0.222 |
| 25239 | Mean L1 in fornix cres+stria terminalis on FA skeleton (left) | -0.010 | 0.049 |
| 25238 | Mean L1 in fornix cres+stria terminalis on FA skeleton (right) | -0.063 | 0.132 |
| 25205 | Mean L1 in fornix on FA skeleton | 0.002 | 0.390 |
| 25202 | Mean L1 in genu of corpus callosum on FA skeleton | 0.121 | 0.237 |
| 25211 | Mean L1 in inferior cerebellar peduncle on FA skeleton (left) | 0.013 | 0.020 |
| 25210 | Mean L1 in inferior cerebellar peduncle on FA skeleton (right) | 0.018 | -0.029 |
| 25209 | Mean L1 in medial lemniscus on FA skeleton (left) | -0.013 | -0.010 |
| 25208 | Mean L1 in medial lemniscus on FA skeleton (right) | 0.028 | -0.016 |
| 25200 | Mean L1 in middle cerebellar peduncle on FA skeleton | -0.100 | -0.010 |
| 25201 | Mean L1 in pontine crossing tract on FA skeleton | 0.071 | -0.040 |
| 25227 | Mean L1 in posterior corona radiata on FA skeleton (left) | 0.244 | 0.280 |
| 25226 | Mean L1 in posterior corona radiata on FA skeleton (right) | 0.230 | 0.328 |
| 25219 | Mean L1 in posterior limb of internal capsule on FA skeleton (left) | 0.026 | 0.168 |
| 25218 | Mean L1 in posterior limb of internal capsule on FA skeleton (right) | 0.005 | 0.204 |
| 25229 | Mean L1 in posterior thalamic radiation on FA skeleton (left) | 0.350 | 0.092 |
| 25228 | Mean L1 in posterior thalamic radiation on FA skeleton (right) | 0.415 | 0.058 |
| 25221 | Mean L1 in retrolenticular part of internal capsule on FA skeleton (left) | 0.043 | 0.128 |
| 25220 | Mean L1 in retrolenticular part of internal capsule on FA skeleton (right) | 0.091 | 0.166 |
| 25231 | Mean L1 in sagittal stratum on FA skeleton (left) | 0.318 | 0.102 |
| 25230 | Mean L1 in sagittal stratum on FA skeleton (right) | 0.294 | 0.088 |
| 25204 | Mean L1 in splenium of corpus callosum on FA skeleton | 0.048 | 0.215 |
| 25213 | Mean L1 in superior cerebellar peduncle on FA skeleton (left) | -0.020 | 0.196 |
| 25212 | Mean L1 in superior cerebellar peduncle on FA skeleton (right) | -0.038 | 0.189 |
| 25225 | Mean L1 in superior corona radiata on FA skeleton (left) | 0.176 | 0.275 |
| 25224 | Mean L1 in superior corona radiata on FA skeleton (right) | 0.190 | 0.271 |
| 25243 | Mean L1 in superior fronto-occipital fasciculus on FA skeleton (left) | 0.033 | 0.293 |
| 25242 | Mean L1 in superior fronto-occipital fasciculus on FA skeleton (right) | 0.098 | 0.269 |
| 25241 | Mean L1 in superior longitudinal fasciculus on FA skeleton (left) | 0.268 | 0.189 |
| 25240 | Mean L1 in superior longitudinal fasciculus on FA skeleton (right) | 0.236 | 0.207 |
| 25247 | Mean L1 in tapetum on FA skeleton (left) | 0.067 | 0.231 |
| 25246 | Mean L1 in tapetum on FA skeleton (right) | 0.144 | 0.270 |
| 25245 | Mean L1 in uncinate fasciculus on FA skeleton (left) | 0.134 | 0.105 |
| 25244 | Mean L1 in uncinate fasciculus on FA skeleton (right) | 0.131 | 0.107 |
| 25271 | Mean L2 in anterior corona radiata on FA skeleton (left) | 0.204 | 0.347 |
| 25270 | Mean L2 in anterior corona radiata on FA skeleton (right) | 0.178 | 0.325 |
| 25265 | Mean L2 in anterior limb of internal capsule on FA skeleton (left) | 0.069 | 0.172 |
| 25264 | Mean L2 in anterior limb of internal capsule on FA skeleton (right) | 0.039 | 0.155 |
| 25251 | Mean L2 in body of corpus callosum on FA skeleton | 0.160 | 0.256 |
| 25263 | Mean L2 in cerebral peduncle on FA skeleton (left) | 0.143 | 0.203 |
| 25262 | Mean L2 in cerebral peduncle on FA skeleton (right) | 0.210 | 0.171 |
| 25283 | Mean L2 in cingulum cingulate gyrus on FA skeleton (left) | 0.128 | 0.225 |
| 25282 | Mean L2 in cingulum cingulate gyrus on FA skeleton (right) | 0.090 | 0.230 |
| 25285 | Mean L2 in cingulum hippocampus on FA skeleton (left) | 0.216 | 0.145 |
| 25284 | Mean L2 in cingulum hippocampus on FA skeleton (right) | 0.266 | 0.092 |
| 25255 | Mean L2 in corticospinal tract on FA skeleton (left) | 0.136 | 0.055 |
| 25254 | Mean L2 in corticospinal tract on FA skeleton (right) | 0.094 | 0.031 |
| 25281 | Mean L2 in external capsule on FA skeleton (left) | 0.098 | 0.349 |
| 25280 | Mean L2 in external capsule on FA skeleton (right) | 0.148 | 0.357 |
| 25287 | Mean L2 in fornix cres+stria terminalis on FA skeleton (left) | 0.026 | 0.352 |
| 25286 | Mean L2 in fornix cres+stria terminalis on FA skeleton (right) | 0.039 | 0.368 |
| 25253 | Mean L2 in fornix on FA skeleton | 0.035 | 0.370 |
| 25250 | Mean L2 in genu of corpus callosum on FA skeleton | 0.101 | 0.275 |
| 25259 | Mean L2 in inferior cerebellar peduncle on FA skeleton (left) | 0.079 | 0.191 |
| 25258 | Mean L2 in inferior cerebellar peduncle on FA skeleton (right) | 0.109 | 0.209 |
| 25257 | Mean L2 in medial lemniscus on FA skeleton (left) | 0.068 | 0.048 |
| 25256 | Mean L2 in medial lemniscus on FA skeleton (right) | 0.058 | 0.052 |
| 25248 | Mean L2 in middle cerebellar peduncle on FA skeleton | 0.172 | 0.090 |
| 25249 | Mean L2 in pontine crossing tract on FA skeleton | 0.044 | -0.002 |
| 25275 | Mean L2 in posterior corona radiata on FA skeleton (left) | 0.201 | 0.189 |
| 25274 | Mean L2 in posterior corona radiata on FA skeleton (right) | 0.227 | 0.155 |
| 25267 | Mean L2 in posterior limb of internal capsule on FA skeleton (left) | -0.011 | 0.041 |
| 25266 | Mean L2 in posterior limb of internal capsule on FA skeleton (right) | 0.040 | 0.066 |
| 25277 | Mean L2 in posterior thalamic radiation on FA skeleton (left) | 0.351 | 0.276 |
| 25276 | Mean L2 in posterior thalamic radiation on FA skeleton (right) | 0.433 | 0.280 |
| 25269 | Mean L2 in retrolenticular part of internal capsule on FA skeleton (left) | 0.056 | 0.149 |
| 25268 | Mean L2 in retrolenticular part of internal capsule on FA skeleton (right) | 0.164 | 0.123 |
| 25279 | Mean L2 in sagittal stratum on FA skeleton (left) | 0.350 | 0.174 |
| 25278 | Mean L2 in sagittal stratum on FA skeleton (right) | 0.356 | 0.181 |
| 25252 | Mean L2 in splenium of corpus callosum on FA skeleton | 0.181 | 0.048 |
| 25261 | Mean L2 in superior cerebellar peduncle on FA skeleton (left) | -0.033 | 0.080 |
| 25260 | Mean L2 in superior cerebellar peduncle on FA skeleton (right) | -0.002 | 0.069 |
| 25273 | Mean L2 in superior corona radiata on FA skeleton (left) | 0.156 | 0.208 |
| 25272 | Mean L2 in superior corona radiata on FA skeleton (right) | 0.186 | 0.212 |
| 25291 | Mean L2 in superior fronto-occipital fasciculus on FA skeleton (left) | 0.038 | 0.307 |
| 25290 | Mean L2 in superior fronto-occipital fasciculus on FA skeleton (right) | 0.120 | 0.306 |
| 25289 | Mean L2 in superior longitudinal fasciculus on FA skeleton (left) | 0.212 | 0.208 |
| 25288 | Mean L2 in superior longitudinal fasciculus on FA skeleton (right) | 0.221 | 0.215 |
| 25295 | Mean L2 in tapetum on FA skeleton (left) | 0.019 | 0.149 |
| 25294 | Mean L2 in tapetum on FA skeleton (right) | 0.177 | 0.225 |
| 25293 | Mean L2 in uncinate fasciculus on FA skeleton (left) | 0.100 | 0.191 |
| 25292 | Mean L2 in uncinate fasciculus on FA skeleton (right) | 0.238 | 0.173 |
| 25319 | Mean L3 in anterior corona radiata on FA skeleton (left) | 0.215 | 0.364 |
| 25318 | Mean L3 in anterior corona radiata on FA skeleton (right) | 0.192 | 0.346 |
| 25313 | Mean L3 in anterior limb of internal capsule on FA skeleton (left) | 0.068 | 0.342 |
| 25312 | Mean L3 in anterior limb of internal capsule on FA skeleton (right) | 0.053 | 0.346 |
| 25299 | Mean L3 in body of corpus callosum on FA skeleton | 0.165 | 0.261 |
| 25311 | Mean L3 in cerebral peduncle on FA skeleton (left) | 0.134 | 0.203 |
| 25310 | Mean L3 in cerebral peduncle on FA skeleton (right) | 0.163 | 0.180 |
| 25331 | Mean L3 in cingulum cingulate gyrus on FA skeleton (left) | 0.214 | 0.215 |
| 25330 | Mean L3 in cingulum cingulate gyrus on FA skeleton (right) | 0.160 | 0.203 |
| 25333 | Mean L3 in cingulum hippocampus on FA skeleton (left) | 0.300 | 0.143 |
| 25332 | Mean L3 in cingulum hippocampus on FA skeleton (right) | 0.319 | 0.132 |
| 25303 | Mean L3 in corticospinal tract on FA skeleton (left) | 0.128 | 0.048 |
| 25302 | Mean L3 in corticospinal tract on FA skeleton (right) | 0.109 | 0.022 |
| 25329 | Mean L3 in external capsule on FA skeleton (left) | 0.113 | 0.305 |
| 25328 | Mean L3 in external capsule on FA skeleton (right) | 0.156 | 0.298 |
| 25335 | Mean L3 in fornix cres+stria terminalis on FA skeleton (left) | 0.018 | 0.297 |
| 25334 | Mean L3 in fornix cres+stria terminalis on FA skeleton (right) | 0.039 | 0.337 |
| 25301 | Mean L3 in fornix on FA skeleton | 0.024 | 0.370 |
| 25298 | Mean L3 in genu of corpus callosum on FA skeleton | 0.122 | 0.326 |
| 25307 | Mean L3 in inferior cerebellar peduncle on FA skeleton (left) | 0.133 | 0.160 |
| 25306 | Mean L3 in inferior cerebellar peduncle on FA skeleton (right) | 0.158 | 0.162 |
| 25305 | Mean L3 in medial lemniscus on FA skeleton (left) | 0.078 | 0.053 |
| 25304 | Mean L3 in medial lemniscus on FA skeleton (right) | 0.081 | 0.049 |
| 25296 | Mean L3 in middle cerebellar peduncle on FA skeleton | 0.177 | 0.128 |
| 25297 | Mean L3 in pontine crossing tract on FA skeleton | 0.112 | 0.021 |
| 25323 | Mean L3 in posterior corona radiata on FA skeleton (left) | 0.234 | 0.271 |
| 25322 | Mean L3 in posterior corona radiata on FA skeleton (right) | 0.228 | 0.287 |
| 25315 | Mean L3 in posterior limb of internal capsule on FA skeleton (left) | 0.090 | 0.209 |
| 25314 | Mean L3 in posterior limb of internal capsule on FA skeleton (right) | 0.104 | 0.220 |
| 25325 | Mean L3 in posterior thalamic radiation on FA skeleton (left) | 0.407 | 0.263 |
| 25324 | Mean L3 in posterior thalamic radiation on FA skeleton (right) | 0.487 | 0.255 |
| 25317 | Mean L3 in retrolenticular part of internal capsule on FA skeleton (left) | 0.094 | 0.161 |
| 25316 | Mean L3 in retrolenticular part of internal capsule on FA skeleton (right) | 0.188 | 0.166 |
| 25327 | Mean L3 in sagittal stratum on FA skeleton (left) | 0.387 | 0.215 |
| 25326 | Mean L3 in sagittal stratum on FA skeleton (right) | 0.398 | 0.206 |
| 25300 | Mean L3 in splenium of corpus callosum on FA skeleton | 0.170 | 0.222 |
| 25309 | Mean L3 in superior cerebellar peduncle on FA skeleton (left) | -0.038 | 0.078 |
| 25308 | Mean L3 in superior cerebellar peduncle on FA skeleton (right) | -0.027 | 0.068 |
| 25321 | Mean L3 in superior corona radiata on FA skeleton (left) | 0.198 | 0.340 |
| 25320 | Mean L3 in superior corona radiata on FA skeleton (right) | 0.235 | 0.328 |
| 25339 | Mean L3 in superior fronto-occipital fasciculus on FA skeleton (left) | 0.071 | 0.330 |
| 25338 | Mean L3 in superior fronto-occipital fasciculus on FA skeleton (right) | 0.166 | 0.327 |
| 25337 | Mean L3 in superior longitudinal fasciculus on FA skeleton (left) | 0.233 | 0.237 |
| 25336 | Mean L3 in superior longitudinal fasciculus on FA skeleton (right) | 0.230 | 0.235 |
| 25343 | Mean L3 in tapetum on FA skeleton (left) | 0.047 | 0.232 |
| 25342 | Mean L3 in tapetum on FA skeleton (right) | 0.177 | 0.277 |
| 25341 | Mean L3 in uncinate fasciculus on FA skeleton (left) | 0.159 | 0.165 |
| 25340 | Mean L3 in uncinate fasciculus on FA skeleton (right) | 0.248 | 0.162 |
| 25127 | Mean MD in anterior corona radiata on FA skeleton (left) | 0.223 | 0.345 |
| 25126 | Mean MD in anterior corona radiata on FA skeleton (right) | 0.213 | 0.338 |
| 25121 | Mean MD in anterior limb of internal capsule on FA skeleton (left) | 0.075 | 0.321 |
| 25120 | Mean MD in anterior limb of internal capsule on FA skeleton (right) | 0.040 | 0.321 |
| 25107 | Mean MD in body of corpus callosum on FA skeleton | 0.174 | 0.285 |
| 25119 | Mean MD in cerebral peduncle on FA skeleton (left) | 0.088 | 0.093 |
| 25118 | Mean MD in cerebral peduncle on FA skeleton (right) | 0.141 | 0.095 |
| 25139 | Mean MD in cingulum cingulate gyrus on FA skeleton (left) | 0.257 | 0.168 |
| 25138 | Mean MD in cingulum cingulate gyrus on FA skeleton (right) | 0.214 | 0.168 |
| 25141 | Mean MD in cingulum hippocampus on FA skeleton (left) | 0.236 | 0.108 |
| 25140 | Mean MD in cingulum hippocampus on FA skeleton (right) | 0.249 | 0.089 |
| 25111 | Mean MD in corticospinal tract on FA skeleton (left) | 0.110 | 0.008 |
| 25110 | Mean MD in corticospinal tract on FA skeleton (right) | 0.077 | -0.010 |
| 25137 | Mean MD in external capsule on FA skeleton (left) | 0.127 | 0.327 |
| 25136 | Mean MD in external capsule on FA skeleton (right) | 0.163 | 0.316 |
| 25143 | Mean MD in fornix cres+stria terminalis on FA skeleton (left) | 0.013 | 0.265 |
| 25142 | Mean MD in fornix cres+stria terminalis on FA skeleton (right) | 0.004 | 0.314 |
| 25109 | Mean MD in fornix on FA skeleton | 0.023 | 0.379 |
| 25106 | Mean MD in genu of corpus callosum on FA skeleton | 0.131 | 0.322 |
| 25115 | Mean MD in inferior cerebellar peduncle on FA skeleton (left) | 0.090 | 0.151 |
| 25114 | Mean MD in inferior cerebellar peduncle on FA skeleton (right) | 0.121 | 0.140 |
| 25113 | Mean MD in medial lemniscus on FA skeleton (left) | 0.040 | 0.026 |
| 25112 | Mean MD in medial lemniscus on FA skeleton (right) | 0.058 | 0.024 |
| 25104 | Mean MD in middle cerebellar peduncle on FA skeleton | 0.085 | 0.073 |
| 25105 | Mean MD in pontine crossing tract on FA skeleton | 0.078 | -0.011 |
| 25131 | Mean MD in posterior corona radiata on FA skeleton (left) | 0.242 | 0.268 |
| 25130 | Mean MD in posterior corona radiata on FA skeleton (right) | 0.248 | 0.286 |
| 25123 | Mean MD in posterior limb of internal capsule on FA skeleton (left) | 0.046 | 0.217 |
| 25122 | Mean MD in posterior limb of internal capsule on FA skeleton (right) | 0.061 | 0.246 |
| 25133 | Mean MD in posterior thalamic radiation on FA skeleton (left) | 0.424 | 0.240 |
| 25132 | Mean MD in posterior thalamic radiation on FA skeleton (right) | 0.516 | 0.227 |
| 25125 | Mean MD in retrolenticular part of internal capsule on FA skeleton (left) | 0.076 | 0.177 |
| 25124 | Mean MD in retrolenticular part of internal capsule on FA skeleton (right) | 0.171 | 0.183 |
| 25135 | Mean MD in sagittal stratum on FA skeleton (left) | 0.425 | 0.195 |
| 25134 | Mean MD in sagittal stratum on FA skeleton (right) | 0.409 | 0.184 |
| 25108 | Mean MD in splenium of corpus callosum on FA skeleton | 0.154 | 0.222 |
| 25117 | Mean MD in superior cerebellar peduncle on FA skeleton (left) | -0.036 | 0.159 |
| 25116 | Mean MD in superior cerebellar peduncle on FA skeleton (right) | -0.029 | 0.145 |
| 25129 | Mean MD in superior corona radiata on FA skeleton (left) | 0.207 | 0.324 |
| 25128 | Mean MD in superior corona radiata on FA skeleton (right) | 0.240 | 0.320 |
| 25147 | Mean MD in superior fronto-occipital fasciculus on FA skeleton (left) | 0.050 | 0.328 |
| 25146 | Mean MD in superior fronto-occipital fasciculus on FA skeleton (right) | 0.139 | 0.329 |
| 25145 | Mean MD in superior longitudinal fasciculus on FA skeleton (left) | 0.280 | 0.249 |
| 25144 | Mean MD in superior longitudinal fasciculus on FA skeleton (right) | 0.264 | 0.253 |
| 25151 | Mean MD in tapetum on FA skeleton (left) | 0.049 | 0.223 |
| 25150 | Mean MD in tapetum on FA skeleton (right) | 0.177 | 0.276 |
| 25149 | Mean MD in uncinate fasciculus on FA skeleton (left) | 0.172 | 0.199 |
| 25148 | Mean MD in uncinate fasciculus on FA skeleton (right) | 0.276 | 0.200 |
| 25175 | Mean MO in anterior corona radiata on FA skeleton (left) | 0.021 | -0.113 |
| 25174 | Mean MO in anterior corona radiata on FA skeleton (right) | 0.023 | -0.091 |
| 25169 | Mean MO in anterior limb of internal capsule on FA skeleton (left) | 0.011 | 0.264 |
| 25168 | Mean MO in anterior limb of internal capsule on FA skeleton (right) | -0.003 | 0.264 |
| 25155 | Mean MO in body of corpus callosum on FA skeleton | -0.029 | -0.284 |
| 25167 | Mean MO in cerebral peduncle on FA skeleton (left) | -0.090 | -0.125 |
| 25166 | Mean MO in cerebral peduncle on FA skeleton (right) | -0.152 | -0.105 |
| 25187 | Mean MO in cingulum cingulate gyrus on FA skeleton (left) | 0.109 | -0.139 |
| 25186 | Mean MO in cingulum cingulate gyrus on FA skeleton (right) | 0.120 | -0.169 |
| 25189 | Mean MO in cingulum hippocampus on FA skeleton (left) | 0.018 | -0.078 |
| 25188 | Mean MO in cingulum hippocampus on FA skeleton (right) | -0.031 | 0.001 |
| 25159 | Mean MO in corticospinal tract on FA skeleton (left) | -0.051 | -0.066 |
| 25158 | Mean MO in corticospinal tract on FA skeleton (right) | -0.054 | -0.067 |
| 25185 | Mean MO in external capsule on FA skeleton (left) | 0.036 | -0.192 |
| 25184 | Mean MO in external capsule on FA skeleton (right) | -0.001 | -0.221 |
| 25191 | Mean MO in fornix cres+stria terminalis on FA skeleton (left) | -0.004 | -0.256 |
| 25190 | Mean MO in fornix cres+stria terminalis on FA skeleton (right) | -0.042 | -0.224 |
| 25157 | Mean MO in fornix on FA skeleton | -0.049 | -0.190 |
| 25154 | Mean MO in genu of corpus callosum on FA skeleton | 0.045 | -0.103 |
| 25163 | Mean MO in inferior cerebellar peduncle on FA skeleton (left) | -0.049 | -0.159 |
| 25162 | Mean MO in inferior cerebellar peduncle on FA skeleton (right) | -0.062 | -0.188 |
| 25161 | Mean MO in medial lemniscus on FA skeleton (left) | 0.008 | -0.054 |
| 25160 | Mean MO in medial lemniscus on FA skeleton (right) | -0.008 | -0.061 |
| 25152 | Mean MO in middle cerebellar peduncle on FA skeleton | -0.187 | -0.027 |
| 25153 | Mean MO in pontine crossing tract on FA skeleton | 0.083 | -0.004 |
| 25179 | Mean MO in posterior corona radiata on FA skeleton (left) | 0.113 | 0.125 |
| 25178 | Mean MO in posterior corona radiata on FA skeleton (right) | 0.006 | 0.211 |
| 25171 | Mean MO in posterior limb of internal capsule on FA skeleton (left) | 0.068 | 0.127 |
| 25170 | Mean MO in posterior limb of internal capsule on FA skeleton (right) | 0.028 | 0.133 |
| 25181 | Mean MO in posterior thalamic radiation on FA skeleton (left) | 0.106 | -0.152 |
| 25180 | Mean MO in posterior thalamic radiation on FA skeleton (right) | 0.082 | -0.169 |
| 25173 | Mean MO in retrolenticular part of internal capsule on FA skeleton (left) | 0.034 | -0.078 |
| 25172 | Mean MO in retrolenticular part of internal capsule on FA skeleton (right) | -0.005 | -0.003 |
| 25183 | Mean MO in sagittal stratum on FA skeleton (left) | 0.048 | -0.046 |
| 25182 | Mean MO in sagittal stratum on FA skeleton (right) | 0.046 | -0.078 |
| 25156 | Mean MO in splenium of corpus callosum on FA skeleton | -0.021 | 0.162 |
| 25165 | Mean MO in superior cerebellar peduncle on FA skeleton (left) | -0.068 | -0.040 |
| 25164 | Mean MO in superior cerebellar peduncle on FA skeleton (right) | -0.099 | -0.016 |
| 25177 | Mean MO in superior corona radiata on FA skeleton (left) | 0.058 | 0.159 |
| 25176 | Mean MO in superior corona radiata on FA skeleton (right) | 0.051 | 0.135 |
| 25195 | Mean MO in superior fronto-occipital fasciculus on FA skeleton (left) | 0.034 | -0.117 |
| 25194 | Mean MO in superior fronto-occipital fasciculus on FA skeleton (right) | -0.002 | -0.081 |
| 25193 | Mean MO in superior longitudinal fasciculus on FA skeleton (left) | -0.004 | -0.021 |
| 25192 | Mean MO in superior longitudinal fasciculus on FA skeleton (right) | -0.021 | -0.017 |
| 25199 | Mean MO in tapetum on FA skeleton (left) | 0.070 | 0.187 |
| 25198 | Mean MO in tapetum on FA skeleton (right) | 0.007 | 0.119 |
| 25197 | Mean MO in uncinate fasciculus on FA skeleton (left) | 0.061 | -0.105 |
| 25196 | Mean MO in uncinate fasciculus on FA skeleton (right) | -0.031 | -0.075 |
| 25415 | Mean OD in anterior corona radiata on FA skeleton (left) | -0.066 | 0.096 |
| 25414 | Mean OD in anterior corona radiata on FA skeleton (right) | -0.093 | 0.037 |
| 25409 | Mean OD in anterior limb of internal capsule on FA skeleton (left) | -0.047 | 0.002 |
| 25408 | Mean OD in anterior limb of internal capsule on FA skeleton (right) | -0.018 | 0.020 |
| 25395 | Mean OD in body of corpus callosum on FA skeleton | 0.044 | 0.135 |
| 25407 | Mean OD in cerebral peduncle on FA skeleton (left) | 0.084 | 0.153 |
| 25406 | Mean OD in cerebral peduncle on FA skeleton (right) | 0.054 | 0.122 |
| 25427 | Mean OD in cingulum cingulate gyrus on FA skeleton (left) | -0.073 | 0.125 |
| 25426 | Mean OD in cingulum cingulate gyrus on FA skeleton (right) | -0.083 | 0.156 |
| 25429 | Mean OD in cingulum hippocampus on FA skeleton (left) | 0.041 | 0.110 |
| 25428 | Mean OD in cingulum hippocampus on FA skeleton (right) | 0.127 | 0.069 |
| 25399 | Mean OD in corticospinal tract on FA skeleton (left) | 0.095 | 0.075 |
| 25398 | Mean OD in corticospinal tract on FA skeleton (right) | 0.059 | 0.085 |
| 25425 | Mean OD in external capsule on FA skeleton (left) | -0.091 | 0.039 |
| 25424 | Mean OD in external capsule on FA skeleton (right) | -0.056 | 0.053 |
| 25431 | Mean OD in fornix cres+stria terminalis on FA skeleton (left) | 0.004 | 0.231 |
| 25430 | Mean OD in fornix cres+stria terminalis on FA skeleton (right) | 0.055 | 0.208 |
| 25397 | Mean OD in fornix on FA skeleton | -0.016 | 0.221 |
| 25394 | Mean OD in genu of corpus callosum on FA skeleton | -0.026 | -0.002 |
| 25403 | Mean OD in inferior cerebellar peduncle on FA skeleton (left) | 0.183 | 0.134 |
| 25402 | Mean OD in inferior cerebellar peduncle on FA skeleton (right) | 0.179 | 0.167 |
| 25401 | Mean OD in medial lemniscus on FA skeleton (left) | 0.108 | 0.089 |
| 25400 | Mean OD in medial lemniscus on FA skeleton (right) | 0.163 | 0.102 |
| 25392 | Mean OD in middle cerebellar peduncle on FA skeleton | 0.290 | 0.112 |
| 25393 | Mean OD in pontine crossing tract on FA skeleton | 0.012 | 0.067 |
| 25419 | Mean OD in posterior corona radiata on FA skeleton (left) | -0.166 | -0.129 |
| 25418 | Mean OD in posterior corona radiata on FA skeleton (right) | -0.131 | -0.213 |
| 25411 | Mean OD in posterior limb of internal capsule on FA skeleton (left) | -0.027 | -0.076 |
| 25410 | Mean OD in posterior limb of internal capsule on FA skeleton (right) | -0.030 | -0.091 |
| 25421 | Mean OD in posterior thalamic radiation on FA skeleton (left) | -0.144 | 0.111 |
| 25420 | Mean OD in posterior thalamic radiation on FA skeleton (right) | -0.159 | 0.145 |
| 25413 | Mean OD in retrolenticular part of internal capsule on FA skeleton (left) | 0.001 | 0.031 |
| 25412 | Mean OD in retrolenticular part of internal capsule on FA skeleton (right) | 0.014 | -0.038 |
| 25423 | Mean OD in sagittal stratum on FA skeleton (left) | -0.126 | 0.056 |
| 25422 | Mean OD in sagittal stratum on FA skeleton (right) | -0.105 | 0.077 |
| 25396 | Mean OD in splenium of corpus callosum on FA skeleton | -0.006 | -0.147 |
| 25405 | Mean OD in superior cerebellar peduncle on FA skeleton (left) | 0.044 | -0.076 |
| 25404 | Mean OD in superior cerebellar peduncle on FA skeleton (right) | 0.042 | -0.085 |
| 25417 | Mean OD in superior corona radiata on FA skeleton (left) | -0.056 | -0.105 |
| 25416 | Mean OD in superior corona radiata on FA skeleton (right) | -0.059 | -0.105 |
| 25435 | Mean OD in superior fronto-occipital fasciculus on FA skeleton (left) | -0.034 | -0.048 |
| 25434 | Mean OD in superior fronto-occipital fasciculus on FA skeleton (right) | 0.042 | -0.035 |
| 25433 | Mean OD in superior longitudinal fasciculus on FA skeleton (left) | -0.090 | 0.029 |
| 25432 | Mean OD in superior longitudinal fasciculus on FA skeleton (right) | -0.050 | 0.024 |
| 25439 | Mean OD in tapetum on FA skeleton (left) | -0.045 | -0.161 |
| 25438 | Mean OD in tapetum on FA skeleton (right) | -0.064 | -0.090 |
| 25437 | Mean OD in uncinate fasciculus on FA skeleton (left) | -0.073 | 0.023 |
| 25436 | Mean OD in uncinate fasciculus on FA skeleton (right) | 0.032 | 0.038 |
